# Causality Analysis in Major Depressive Disorder for Early Prediction of Treatment Outcomes with Pharmacological and Neuromodulation Therapies

**DOI:** 10.1101/2025.05.30.25328650

**Authors:** Madhurima Bhattacharjee, Ioannis Vlachos, Aditi Kathpalia, Jaroslav Hlinka, Martin Brunovský, Martin Bareš, Milan Paluš

## Abstract

**Objective:** Major depressive disorder affects millions of people globally, which lacks dependable biomarkers for the early identification of treatment success. This study examines two treatment modalities, pharmacological and neurostimulation. The aim is to discern alterations in brain connection patterns and direction of influence among various regions during the initial phase of the two treatment approaches.

**Approach:** We perform an information theory-based causality analysis on instantaneous phase time series data derived from electroencephalography recordings of 176 patients who underwent the aforementioned treatments. Each patient was recorded twice: prior to the commencement of treatment (visit 1) and one week after the initiation of treatment (visit 2).

**Main results:** Two independent outcomes were observed. Initially, we discovered that different treatment modalities have different impacts on the brain’s connectivity and the direction of influence during the course of one week of treatment. The cohort administered with pharmacological agents exhibited a notable increase in both global and local information transmission in the brain within the β_2_ (18Hz - 25.5Hz) frequency range, whereas the group subjected to stimulation exhibited a notable increase within the δ (1.5Hz - 3.5Hz) band. Secondly, potential respondents and nonrespondents to pharmacological therapy had distinct baseline brain causality effects at the initial session, with individuals who reacted to pharmacological therapy having considerably reduced information flow strength in the α (8Hz - 12Hz) band compared to those who did not respond to treatment.

**Significance:** Causality analysis is conducted on the phases of the data, yielding more accurate conclusions by minimising the possible noise introduced by the signal amplitude. Our study provides significant insights into the directional influence inside the brain during depression and subsequent treatment.

## 1 Introduction

Major depressive disorder (MDD) is a widespread mental health condition affecting millions of people worldwide [1]. It can contribute to various physiological illnesses characterised by enduring unhappiness, loss of interest in activities, and a range of physical and cognitive symptoms. Despite the development of novel treatment strategies for MDD, there remains an absence of reliable biomarkers to assess illness progression and therapy efficacy. This deficit is partially attributable to the diagnostic criteria for MDD, which predominantly focus on behavioural aspects and depend on self-reported symptoms from patients. There is an urgent necessity for novel biomarkers of MDD to facilitate the development of revolutionary therapeutics and improve existing treatments. Determining efficient, non-invasive diagnostic and therapeutic approaches for MDD poses a significant challenge in clinical neuroscience.

Electroencephalography (EEG) is a widely used, non-invasive neuroimaging technique that measures cerebral electrical activity with a high temporal resolution in the millisecond range, providing benefits over alternative brain imaging modalities. Numerous studies have examined changes in brain functional connectivity, i.e., the pattern of statistical dependencies between remote neurophysiological processes [2], in individuals with MDD, utilising EEG data [3]. EEG functional connectivity between brain areas is generally assessed by correlation or coherence estimates [4, 5, 6] derived from neural signals captured by several electrodes. Band power analysis across different EEG frequency bands is another common approach for extracting quantitative EEG markers [7]. Additionally, researchers have used other feature extraction methods such as graph theory [8, 9], dynamic causal modelling [10], machine learning [11, 12], deep learning [13, 14], and its advanced variants such as artificial intelligence instead of classical statistical analysis.

However, to date, little research has been conducted on information theory-based causality analysis in the brains of individuals with MDD using EEG data [15, 16, 17, 18]. Causality analysis goes beyond correlation or coherence to identify the cause-and-effect relationships between variables. The information-theoretic generalisation of the Granger causality concept [19] provides a solid mathematical basis for several reliable causality inference methods [20, 21], including the phase dynamics adaptation used in this study [22]. Analysing causality in multichannel EEG signals might enhance the formulation of more effective treatment strategies by elucidating the variations in the direction and intensity of causal influences among the various localised processes captured by EEG channels, thereby illuminating the brain’s fundamental mechanisms.

In this study, we perform a multivariate causality analysis on EEG data recorded from participants diagnosed with MDD and treated with medication or neurostimulation. In addition to understanding the changes in directional connectivity in the brain resulting from treatment, it is crucial to ascertain whether various treatment modalities influence the brain systems differently. This might be particularly advantageous in the early phase of treatment, as it may facilitate the prompt identification of traits that can predict the clinical outcome, which is here assessed after 4-6 weeks through patient self-reporting.

To estimate causality across brain regions, we employ an information-theoretic method called partial mutual information from mixed embedding (PMIME) [23, 22], and examine alterations in brain connectivity and the directional influence among these regions following the initiation of antidepressant treatment. PMIME is a model-free method that is widely applicable for identifying direct causal links between two components in multivariate systems, such as multi-channel EEG signals. This study employs pPMIME, which is a version of PMIME that operates on instantaneous phase time series of EEG data. Phase-based causality analysis effectively reveals genuine causal relationships in weakly linked oscillatory systems, mitigating the effect of noise potentially introduced by amplitude variations. It is a well-known fact that weakly coupled oscillators can alter the phases of each other while their amplitudes remain unchanged [24]. We leverage the oscillatory nature of EEG signals and employ phase-based causality analysis to achieve more reliable results. In Section 2 we present the methodology and data used in the analysis. In Section 3 we present the obtained results and then discuss and conclude them in Sections 4 and 5.

## 2 Methods and Materials

### 2.1 Phase-based causality analysis (pPMIME)

PMIME/pPMIME [22] is an iterative method that uses forward selection to estimate inherent lagged dependence in coupled multivariate nonlinear dynamical systems. It generates mixed embedding vectors from multiple observed time series of connected subsystems to reveal the causal relationships. The method employs mutual information/conditional mutual information (MI/CMI) *I*(*X*;*Y*)/*I*(*X*;*Y* | *Z*) as a selection criterion and uses a statistical test as a stopping criterion of the forward selection procedure. MI measures the average shared information between random variables *X* and *Y*, whereas CMI measures the same but conditioned by variable *Z*. Consider a coupled dynamical system consisting of *K* subsystems. The time series data for these *K* variables are denoted by *x*_*i,t*_, where *t* = 1, …, *N* is the time index and *i* = 1, …, *K* is the index of the variables. PMIME aims to predict the *T*-steps ahead future values of a target variable *x*_*i*_ by finding a mixed embedding vector **w**_*i*_ = [*w*_(1)_, *w*_(2)_ …] with an undetermined dimension that maximises the MI term *I* (*x*_*i,t*+*T*_; **w**_*i*_). The elements of **w**_*i*_ belong to the superset 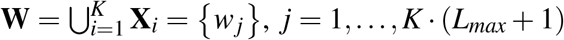, with the sets **X**_*i*_ containing the lagged version of each variable, i.e. **X**_*i*_ = {*x*_*i, t*_, *x*_*i, t*−1_,…, *x*_*i, t*−*L*_^max^}, *i* = 1, …, *K*, where *L*_*max*_ is the maximum lag, which can be arbitrarily chosen to be appropriately large. Starting from an empty vector 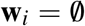, the embedding vector is populated sequentially using a forward selection procedure based on the maximisation of MI/CMI. The CMI terms for step *s* of the procedure are of the form 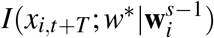, where 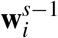 is the embedding vector at the previous step (*s* − 1) and *w*^*^ is any element of **W**. The *w*^*^ that maximises the CMI term is selected to be added in 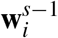 in order to create the new **w**^*s*^. The whole procedure is terminated using a bootstrap resampling test as a stopping criterion, i.e., testing the null hypothesis 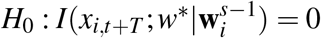. Detailed information regarding the entire procedure can be found in [22, 25]. If for a given *I* (*x*_*i,t*+*T*_; **w**_*i*_), any element of **X** _*j*_ is contained in **w**_*i*_, then we can interpret that *x* _*j*_ causes *x*_*i*_. A causality measure can be derived from the following normalised CMI term, bounded between 0 and 1:

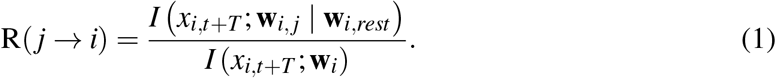

In (1), **w**_*i*_ is split into elements that belong to **X** _*j*_ and all other elements, i.e., **w**_*i*_ = **w**_*i, j*_, **w**_*i,rest*_ and R(*j* → *i*) corresponds to the proportion of information in *x*_*i*_ which can be explained by the past values of *x* _*j*_ when taking into account the values of the rest of *x*_*i*_. PMIME essentially tries to detect which of the present and past values (lags) of *x* _*j*_, *j* = 1 … *K*, i.e., *x* _*j,t*_, *x* _*j,t*−1_ … *x* _*j,t*−*Lmax*_ can be used to optimally predict the future values *x*_*i,t*+*T*_, where T can be any value of *T* = 1, 2, …, *T*_*max*_.

Similarly, when applied to phase time series (*ϕ*_*i,t*_), the algorithm (pPMIME in this case) tries to detect which of the present and past lags of *ϕ*_*j*_, i.e., *ϕ*_*j,t*_, *ϕ*_*j,t*−1_, …, *ϕ*_*j,t*−*Lmax*_ for all *j*, can be used to predict the values of the phase increments Δ_*T*_ *ϕ*_*i,t*+*T*_ = *ϕ*_*i,t*+*T*_ − *ϕ*_*i,t*_. Paluš and Stefanovska demonstrated that, in the bivariate case, Δ_*T*_ *ϕ*_*i,t*+*T*_ yields more sensitive causality tests than *ϕ*_*i,t*+*T*_ itself [26]. Equation (1) is modified accordingly using the phase variables.

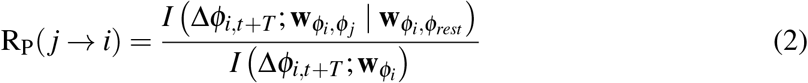

The phase time series *ϕ*_*i,t*_ can be extracted from the original time series *x*_*i,t*_ using various methodologies, but herein we employ the commonly used Hilbert transform (HT) method [27, 28]. Recently, it was shown that for systems with well-behaving phases or narrowly bandpass filtered data (like the EEG filtered in EEG bands), values of *L*_*max*_ = 0 and *T* = 1 are sufficient to capture causality with reduced computational time [25], so this is adopted herein. With respect to the original pPMIME, we make another simple but intuitive logical change. Instead of starting from an empty embedding vector, we start with one that includes the 0^th^ lag of the target variable; i.e., if we are creating embedding vectors for Δ*ϕ*_*i,t*+1_ = *ϕ*_*i,t*+1_ − *ϕ*_*i,t*_, then the embedding vector is initialised as **w**_*i*_ = [*ϕ*_*i,t*_].

### 2.2 Artificial data and method benchmark

We benchmark the pPMIME method for three cases of basic coupled nonlinear oscillators. We study coupled Rössler systems [29, 30] given by sets of the following equations (one set for each oscillator):

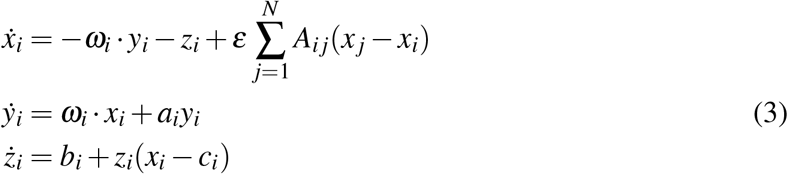

where *i* = 1, …, *N*, and *N* is the total number of subsystems or nodes. The parameters *a* = 0.15, *b* = 0.2, and *c* = 10 were kept constant for all *N* and were chosen so that all subsystems maintain a stable periodic orbit. The elements of the binary (0*/*1) adjacency matrix **A** are denoted by

*A*_*i j*_ and define the connectivity structure of the system, with a value of 1 for *A*_*i j*_ indicating a causal connection from subsystem *j* to *i*. The parameters *ω*_*i*_ determine the main frequency of oscillation of the *i*^th^ subsystem and are set to different values for the subsystems. Finally *ε* is the coupling strength, which we consider to change globally (i.e., it is the same for all subsystems). We studied three cases of 3, 5, and 19 coupled subsystems to validate our method on multi- coupled systems with known directions of influence to imitate real-life scenarios where there are large coupled systems. The values of *ω*_*i*_ were chosen as *ω*_*i*_ ∈ {0.95, 1.00, 1.05} for *N* = 3 (step of 0.05), *ω*_*i*_ ∈ {0.90, 0.95,…, 1.10} for *N* = 5 (step of 0.05), and *ω*_*i*_ ∈ {0.865, 0.880,…, 1.135} for *N* = 19 (step of 0.015). For *N* = 3 and 5, we chose the elements of the adjacency matrix such that each node was unidirectionally coupled, forming a serial connection for *N* = 3 and a “ring” of connections for *N* = 5. For *N* = 19, we generated an adjacency matrix randomly such that only 10% of the connections were present. The adjacency matrices for the three cases are displayed in panels (a), (b), and (c) of Figure 1.

**Fig. 1.**
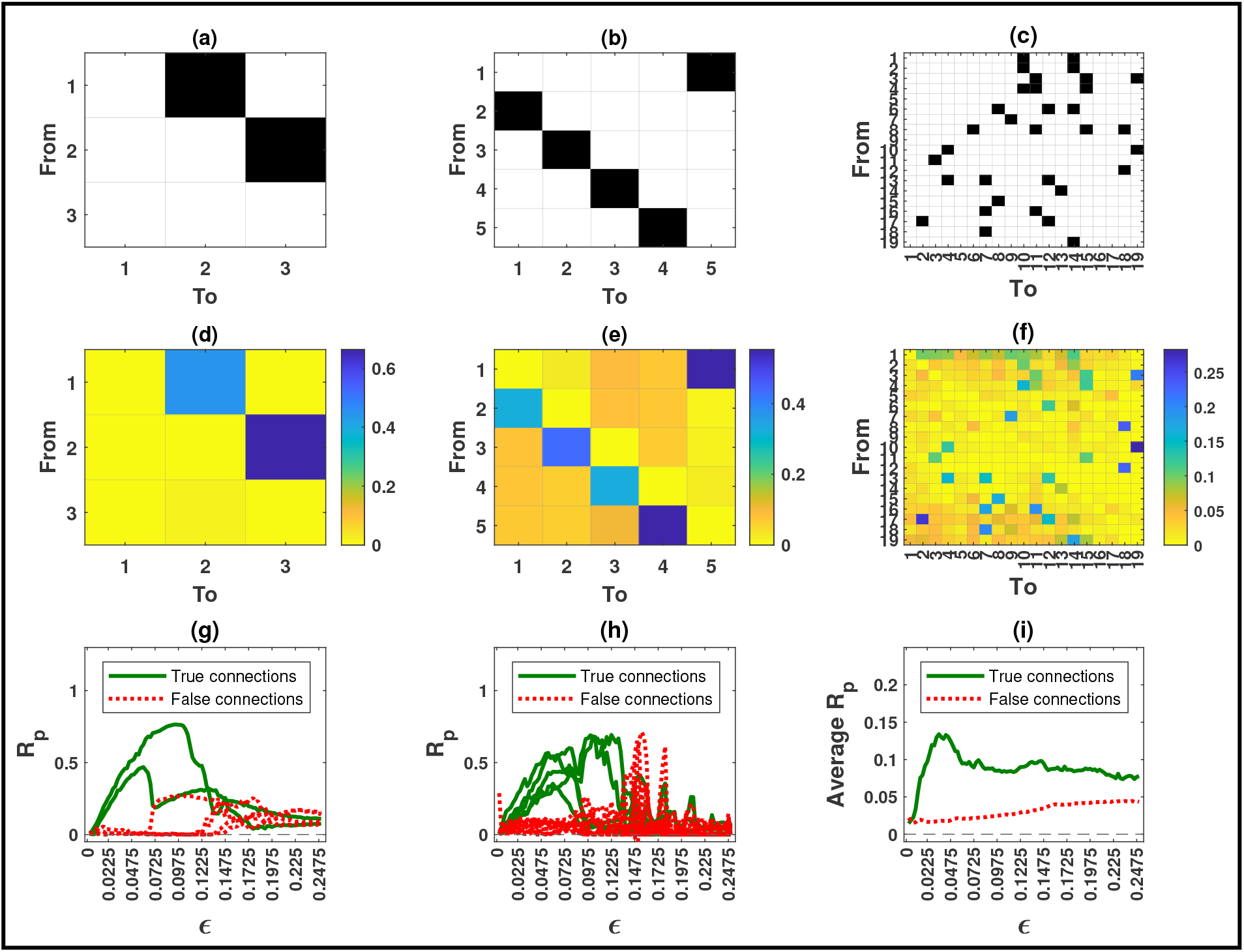
The adjacency matrices for the three cases of coupled Rössler oscillators with *N* = 3, 5 and 19 subsystems in (a), (b), and (c), respectively, and the corresponding pPMIME causality matrices (R_P_ values) for the coupling strength where the average of all real connections was maximum in (d), (e), and (f). R_P_ values as a function of coupling strength for the three networks in (g), (h), and (i). R_P_ values for real connections are plotted in solid green lines and those for non-connections in red dotted lines. For clarity, in panel (i), instead of the individual lines, their averages are plotted.

We studied 100 different values of coupling strength *ε* ∈ {0, 0.0025,…, 0.2475} (step size of 0.0025), and for each one we generated 131072 data points from (3) using the Runge-Kutta (4,5) method (ode45 in MATLAB ver. R2021a). We chose a sampling time of 0.314 time units, and the first 1000 transient points were discarded. The data were Hilbert transformed, and the first and last 2032 time points were discarded to eliminate any distortion of the data owing to the edge effect of the transform. The remaining data were then divided into 31 segments, with each segment containing 4064 time points. pPMIME was then applied to each of these segments, which resulted in 31 (*N*× *N*) causality matrices that contained the R_P_ values for all pairs of *i, j*. The matrices were then averaged over all 31 segments. Each element in the matrix quantifies the causal influence of a particular node on another node, with the diagonal elements corresponding to the self-influences. For example, the (*i, j*)^th^ component of the matrix provides the causal influence of node *i* on node *j*. Panels (d), (e), and (f) in Figure 1 show causality matrices for the three cases of *N*, respectively, each for a selected coupling strength for which the average of all real connections was maximum. The specific values of epsilon for the three cases (*ε* = 0.06, 0.0625, 0.0325 for *N* = 3, 5, and 19, respectively) are small enough to not have widespread synchronisation but large enough so that the obtained R_P_ values are substantial and illustrate the results sufficiently well. As expected, the causality matrices correlate well with the adjacency matrices. Panels (g) and (h) show the R_P_ values for the individual connections for *N* = 3 and *N* = 5, as a function of coupling strength. For the case of *N* = 19, in the panel (i), we show the average of the R_P_ values corresponding to real connections and of those that correspond to no connections, again as a function of coupling, for reasons of visual clarity. The real connections are plotted as solid green lines, and the rest are indicated by red dotted lines. For all three cases and for low coupling strength values (i.e., prior to the synchronisation of the subsystems), the green lines exhibit monotonically increasing trends and exceed the values of the red lines. This indicates that the method can accurately capture the direction of causality between the subsystems. Upon synchronisation of subsystems, causality cannot be uniquely inferred, and we can see that the red lines increase and the green lines decrease. In addition, as the number of subsystems increases, the strength of the detected connections decreases.

### 2.3 EEG data

A total of 176 subjects diagnosed with MDD were analysed for this study. Detailed information regarding the sample and recruitment criteria can be found in previous clinical analysis reports [31, 32, 33, 34]. Details regarding sex, age, and response to antidepressant treatments are shown in Table 1. The patients were subjected to either pharmacological or neurostimulation treatments for four weeks. Individuals who showed a ≥ 50% reduction in the Montgomery-Asberg Depression Rating Scale (MADRS) score [35, 36] were considered as responders to the treatment. Pharmacological treatment includes the following classes of antidepressants: serotonin norepinephrine reuptake inhibitors (SNRI), selective serotonin reuptake inhibitors (SSRI), norepinephrine-dopamine reuptake inhibitors (NDRI), noradrenergic and specific serotonergic antidepressants (NaSSA), and tricyclic antidepressants (TCA). Neurostimulation treatment included repetitive transcranial magnetic stimulation (rTMS) and transcranial direct current stimulation (tDCS). There were two EEG recording sessions for each patient, one before treatment initiation (visit 1) and a second after one week of treatment (visit 2). Data were recorded with the participants lying in a semirecumbent position with their eyes closed in a maximally alert state. Approximately 10 minutes of data were sampled at 250 Hz or 1000 Hz from 19 electrode channels (Fp1, Fp2, F3, F4, C3, C4, P3, P4, O1, O2, F7, F8, T3, T4, T5, T6, Fz, Cz, and Pz) placed according to the International 10-20 system [37].

**Table 1.** Patient demographics and information.

| Attribute | Group | No of patients | Mean ± SD / Percentage |
| --- | --- | --- | --- |
| Age | All | 176 | 45.7 ± 11.32 |
| | Male | 48 | 45.02 ± 13.5 |
| | Female | 128 | 46.07 ± 10.4 |
| SEX | Male | 48 | 27% |
|  | Female | 128 | 73% |
| Therapy | Pharmacological Respondents | 68 | 38.6% |
|  | Pharmacological Nonrespondents | 61 | 34.6% |
|  | Neurostimulation Respondents | 16 | 9.1% |
|  | Neurostimulation Nonrespondents | 31 | 17.7% |

#### 2.3.1 Preprocessing and causality estimation

The data have been used in previous studies and were preprocessed as described in Ref. [38]. In short, the preprocessing included removing extra channels, resampling records to a common 250 Hz sampling frequency, discarding the first and last 30 seconds of each record, re-referencing to the average reference, bandpass filtering the data between 1 and 40 Hz, and removal of data segments with high-power artefacts. The overall aim is to increase the signal-to-noise ratio and make the data more interpretable. More details on the specifics of the preprocessing steps and their individual effects are provided in [38]. We further filtered the data into six different frequency bands to focus on causality related to the specific brain activity in each of these bands. The bands were divided as follows: *δ* : (1.5Hz - 3.5Hz), *θ* : (4Hz - 8Hz), *α*: (8Hz - 12Hz), *β*_1_: (12.5Hz - 17.5Hz), *β*_2_: (18Hz - 25.5Hz) and *γ*: (26Hz - 40Hz). We implement pPMIME on data segments of 8s duration (2000 time points), as the estimation of CMI has large data requirements. Thus, for each data segment, we estimated six 19 × 19 matrices, one for each band. For each subject, a varying number of segments were available, ranging from 14 to 92. The findings of the *γ* band are not used herein because of R_P_ estimation problems due to improper phase extraction at high frequencies with HT (attributed to the extensive range of the *γ* band).

### 2.4 Flow Metrics

The pPMIME method generates causality matrices of dimensions (19 × 19) for the 19 EEG channels. Each element in the matrix quantifies the causal influence of one channel on another. The causality matrices are structured such that the initial eight nodes correspond to channels in the left hemisphere (Fp1, F3, C3, P3, O1, F7, T3, and T5), the subsequent eight nodes correspond to channels in the right hemisphere (Fp2, F4, C4, P4, O2, F8, T4, and T6), and the final three nodes correspond to channels located along the midline of the head (Fz, Cz, and Pz).

We define FLOW metrics based on the average of specific parts from a standard causality matrix, as illustrated in Figure 2. This allows for the quantification of information flow between brain regions derived from the causal relationships identified in the EEG channel data. We define nine FLOW metrics: The Total Flow (*TF*) metric is the aggregate information flow across the 19 channels, calculated by averaging all elements of the matrix. We define 4 metrics for the left hemisphere, *LL, LR, out*_*L*, and *in*_*L*, which respectively represent the information flow from the left hemisphere to itself, to the right hemisphere, to all the regions of the brain, and the information flow from all regions of the brain to it. Similarly, we define metrics for the right hemisphere, which we denote as *RR, RL, out*_*R*, and *in*_*R*. The averages are calculated for the regions indicated by the red boxes labelled **LL, LR, out L, in L, RR, RL, out R**, and **in R** in Figure 2. Additionally, we study the inflow and outflow for each channel separately, which are defined as 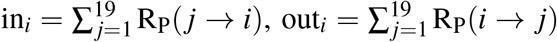, ∀*i* ∈ {1, 2, …, 19}. So in total, we have 47 metrics. The first 9 are the FLOW metrics, which are global and quantify the information flow of the brain as a whole or per brain hemisphere. The other 38 are the INFLOW and OUTFLOW per channel, which are local metrics and characterise small brain regions that correspond to the positions of the electrodes.

**Fig. 2.**
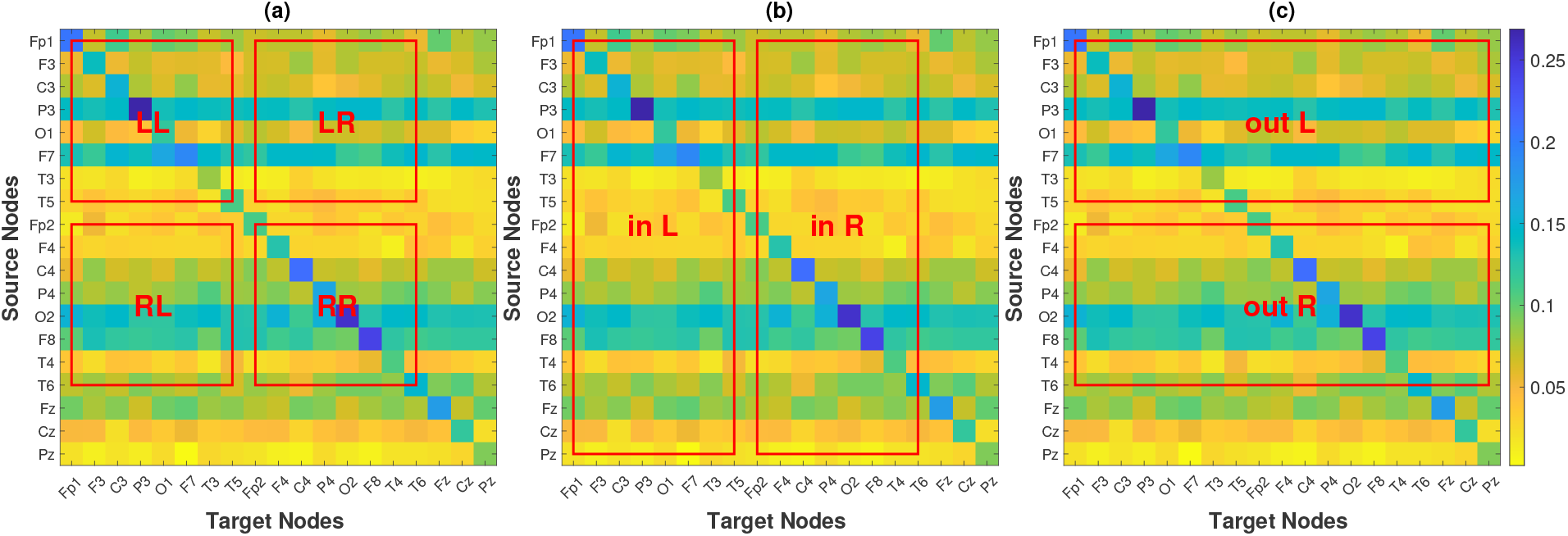
Causality matrix of a subject, along with the regions that are used to define FLOW metrics.

### 2.5 Statistical Analysis

After obtaining the causality matrices for each subject over all time segments, visits, and frequency bands, we compute the metrics defined in Section 2.4 for every time segment of each participant. Subsequently, we calculate the mean of the metrics across all the time segments. Thus, for each band and each of the 47 metrics, we have two sets of 176 values (one set for each visit). Since there are multiple factors (response, treatment, and visit) and one of them is a repeated factor (the EEG recording is performed twice on the same patient), we analysed the data with repeated measures analysis of variance (ANOVA) [39]. The repeated measures ANOVA design includes two independent between-subject factors (response and treatment) and one repeated (dependent) within-subject factor (the visit), each with two levels. The response factor has two levels: respondents and nonrespondents, while the levels for the treatment factor are pharmacological and neurostimulation. For the visit, the two levels are visit 1 and visit 2. The setup of the repeated measures ANOVA model includes the individual factors along with all their interactions (3 pairwise and 1 with all three). The analysis for each EEG frequency band is performed separately. Because of the multiple factors and the “rigidity” of the ANOVA design, we do not actually derive statistical conclusions from these p-values; rather, we utilise these results as preliminary information to guide subsequent analysis. Informed by the outcomes derived from the repeated measures ANOVA study, further post-hoc analysis for the comparison of individual patient subgroups is performed with Student’s t-tests (independent or pairwise), and the effect size of any change/difference is estimated by Cohen’s *d* [40]. Non-parametric tests [41] gave similar results to the t-tests, with p-values generally slightly higher with certain differences in statistical significance for some metrics, but no substantial change in the overall conclusions. The ultimate goal of this analysis is to reveal which, if any, of these metrics has the potential to serve as biomarkers.

Our study is exploratory in nature. There are no specific hypotheses tested where there is an expected difference based on some prior knowledge. What we are doing is a “blind” investigation on which metrics show a difference, and to do this, we employ a comprehensive set of metrics on all possible EEG bands, expecting that most of them will probably not provide statistically significant results. As a matter of fact, we expect our significant results to be constrained to one or, at most, two frequency bands. Furthermore, certain metrics are, by construction, correlated. For example, *out*_*L* (outflow from the left hemisphere) is related to *LL* and *LR* (flow from the left to left and from the left to right hemispheres, respectively); therefore, if one of the latter shows something significant, we expect the former to also show it. The average (absolute) correlation for the FLOW metrics is 0.554, whereas if they were uncorrelated, it should be around 0.043. Because of the high correlation and large number of metrics, global corrections for multiple comparisons are very likely to lead to most p-values being “over-corrected” and rendered insignificant. Since the study is exploratory, over-correcting the p-values is antithetical to the main objective. It has recently been argued that p-value correction should be performed only for the omnibus hypothesis (i.e., experiment-wise) but not always used for statistical claims for individual hypotheses [42, 43]. Of course even in exploratory studies there is a need for some form of correction for individual p-values, but it should be done carefully and tailored to the study; otherwise, there can be severe loss of statistical power [42, 44]. Herein we define three “families” of statistical hypothesis tests based on our three types of metrics (global, inflow, outflow) and use the Benjamini–Yekutieli FDR controlling procedure [45] to obtain adjusted p-values per metric family and EEG band. All p-values in the following section are FDR adjusted and assessed at the *α* = 0.05 significance level. The original, unadjusted p-values from the t-tests and the p-values from the non-parametric rank-based tests are provided in the supplementary materials, Tables 4-7.

## 3 Results

### 3.1 Results from repeated measures ANOVA

The results of the repeated measures ANOVA are graphically presented in Figure 3 and summarised in Table 2. The figure displays a colormap highlighting the p-values for the factors and interactions with *p <* 0.05 for each metric. The colormap is organised vertically in seven blocks separated by red horizontal lines. Each block corresponds to a factor or interaction of the factors, and within the block are the five frequency bands. The horizontal axis displays the metrics. The three families of metrics are separated by black vertical lines. The p-values ≥ 0.05 are all set to yellow, while the colder colours correspond to lower p-values. The text around the colormap denotes factors, bands, and metrics. For example, for the TF metric, the treatment factor is significant in the *δ* band (*p* = 0.012) and is presented in the first column, the first value of the second block. For the same metric, the only other p-values that are below 0.05 are the response-treatment and treatment-visit interactions in the *β*_2_ (*p* = 0.038, *p* = 0.0001, respectively). Table 2 summarises the repeated measures ANOVA results by presenting the number of metrics/bands that provide a significant p-value for each factor/interaction.

**Table 2.**
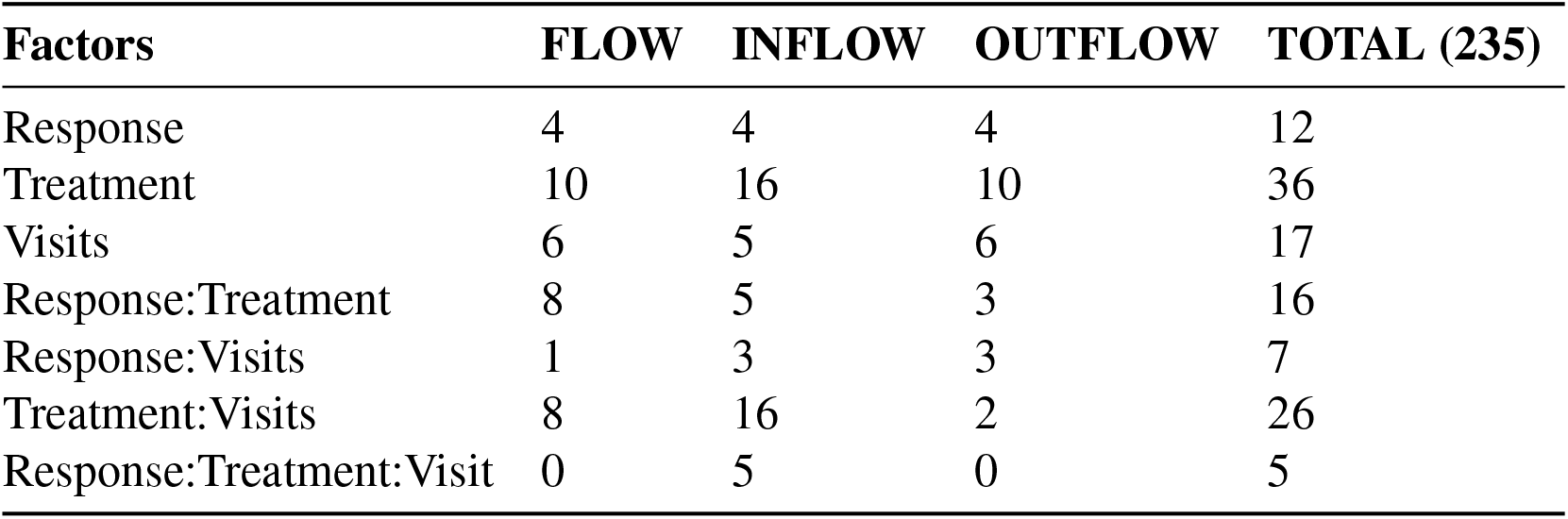
Total number of metrics/band combinations that provide significant p-values for different factors and their interactions.

| Factors | FLOW | INFLOW | OUTFLOW | TOTAL (235) |
| --- | --- | --- | --- | --- |
| Response | 4 | 4 | 4 | 12 |
| Treatment | 10 | 16 | 10 | 36 |
| Visits | 6 | 5 | 6 | 17 |
| Response:Treatment | 8 | 5 | 3 | 16 |
| Response:Visits | 1 | 3 | 3 | 7 |
| Treatment:Visits | 8 | 16 | 2 | 26 |
| Response:Treatment:Visit | 0 | 5 | 0 | 5 |

**Fig. 3.**
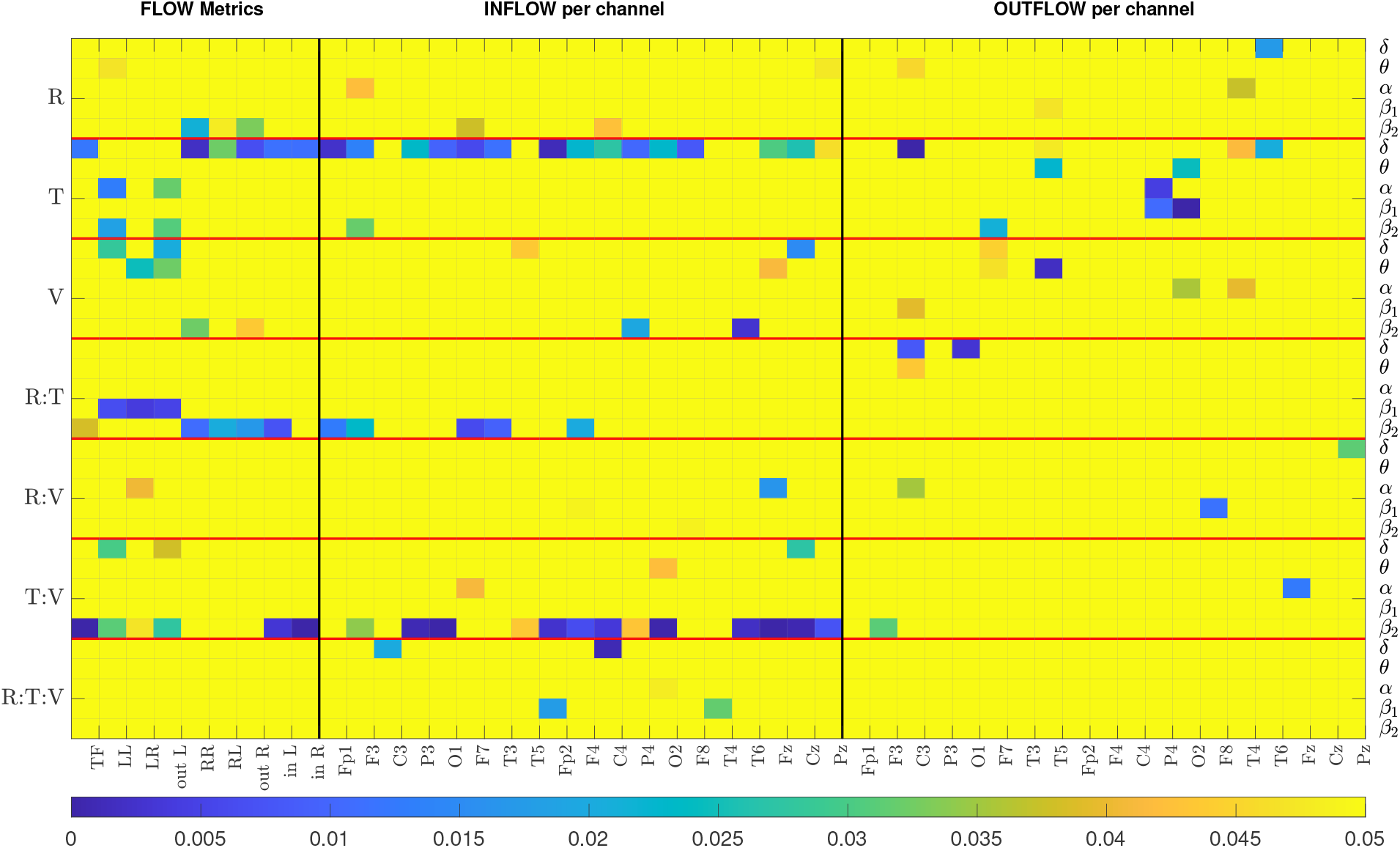
The colormap displays p-values from the repeated measures ANOVA. All p-values above 0.05 are highlighted in yellow. R: Response, T: Treatment, V: Visit. Each row represents the results of a particular band. There are seven blocks corresponding to each factor, separated by a horizontal red line. The metrics corresponding to FLOW, INFLOW and OUTFLOW per channel are separated by a black vertical line.

In Figure 3, we observe that the majority of p-values that are less than 0.05 are mostly for the *δ* and the *β*_2_ bands, and that INFLOW metrics show more cases of statistical significance than the OUTFLOW metrics. The summarised repeated measures ANOVA results from Table 2 show that the three-way interaction factor is not significant for the majority of the metrics and nearly all bands. Only the INFLOW exhibits significant p-values for this interaction for just five metrics spread across all bands. The pairwise interactions between the treatment factor with response and the treatment factor with visit yield the highest number of significant p-values (16 and 26, respectively). On its own, the treatment factor yields 36 statistically significant metric/band combinations. These results indicate that the treatment has the most significant effect on the values of the metrics. Thus, the results suggest analysing the data from the two treatments independently. The single factors of response and visit give significant values for 12 and 17 cases, but their interaction gives 7, and most of them have relatively high p-values (5 out of the 7 are around 0.04 and the 2 around 0.015). This indicates that even though there is a change in the metric values between visits and there exists a difference between those that responded to treatment and those that did not, these two factors are relatively independent. That is, changes in metric values between visits 1 and 2 are not much related to treatment outcome (response). Combining this with the result for the treatment and visit interaction, we can deduce that treatment may be a strong confounding factor when studying the relationship between response to treatment and metric values. We note that since visit 2 was one week after visit 1, but the respondent/nonrespondent assessment occurred at week 4, it is possible that the week 2 metrics (i.e., brain connectivity) may not have changed enough to “capture” the treatment outcome.

Based on the results of the repeated measures ANOVA, we chose to examine the subjects who received pharmacological treatment and neurostimulation independently. Additionally, we perform two distinct analyses. First, we compare the metrics of the two visits. Here we are not concerned with the treatment outcome but with what effect, if any, the two treatments have on the metrics and consequently on the causal relation in the brain. The second analysis compares the metrics between respondents and nonrespondents to treatment. In this case we compare the metrics of visit 1 for the two response groups. We exclude the visit 2 data for two reasons. First, according to the repeated measures ANOVA results, there is no significant interaction between the response and visit interactions. Second, we cannot be sure about the therapeutic state of the patients at visit 2. It is possible that for some subjects the treatment has produced a change in brain connectivity related to the response to therapy, while in others it may not have. Furthermore, we only present the results for the FLOW and INFLOW metrics. The ANOVA results for the OUTFLOW metrics show that the statistically significant results for the cases of interest are either much fewer than the INFLOW metrics (2 versus 16 for the treatment/visit interaction) or spread across bands (e.g., compare Figure 3 results for INFLOW and OUTFLOW for the treatment factor). For OUTFLOW, a minimum of one and a maximum of four channels were sporadically significant in a particular band, whereas all other channels had high p-values. All numerical values of the repeated measures ANOVA p-values are presented in the supplementary materials (Tables 1-3) for the sake of completeness.

#### 3.3.1. Effect of treatment on brain causality

This section presents the results of the change in metrics between visits 1 and 2 for the two therapy groups independently. We perform Student’s t-test (paired) to compare the metric values from two visits. The p-values for all FLOW metrics and all five bands are listed in Table 3. The *β*_2_ and the *δ* bands are primarily significant for the FLOW metrics in the pharmacological and neurostimulation groups, respectively. In the first group, the p-values in the *β*_2_ band indicate a substantial difference between visits across all FLOW metrics. In the second treatment group *TF, LL, LR, out*_*L, in*_*L*, and *in*_*R* are significantly different in the *δ* band. Figure 4 illustrates the variation in all the FLOW metrics between the two visits for the pharmacological group in the *β*_2_ band, and Figure 5 presents the same for the neurostimulation group in the *δ*. No metric achieved statistical significance after FDR correction in the other three frequency bands; therefore, we omit the presentation of these results graphically (figures corresponding to all metrics and all bands are presented in the supplementary materials, Figures 1-5). In both figures, the y-axis of the panels displays the metric values averaged over all subjects along with the standard errors of the mean.

**Table 3.**
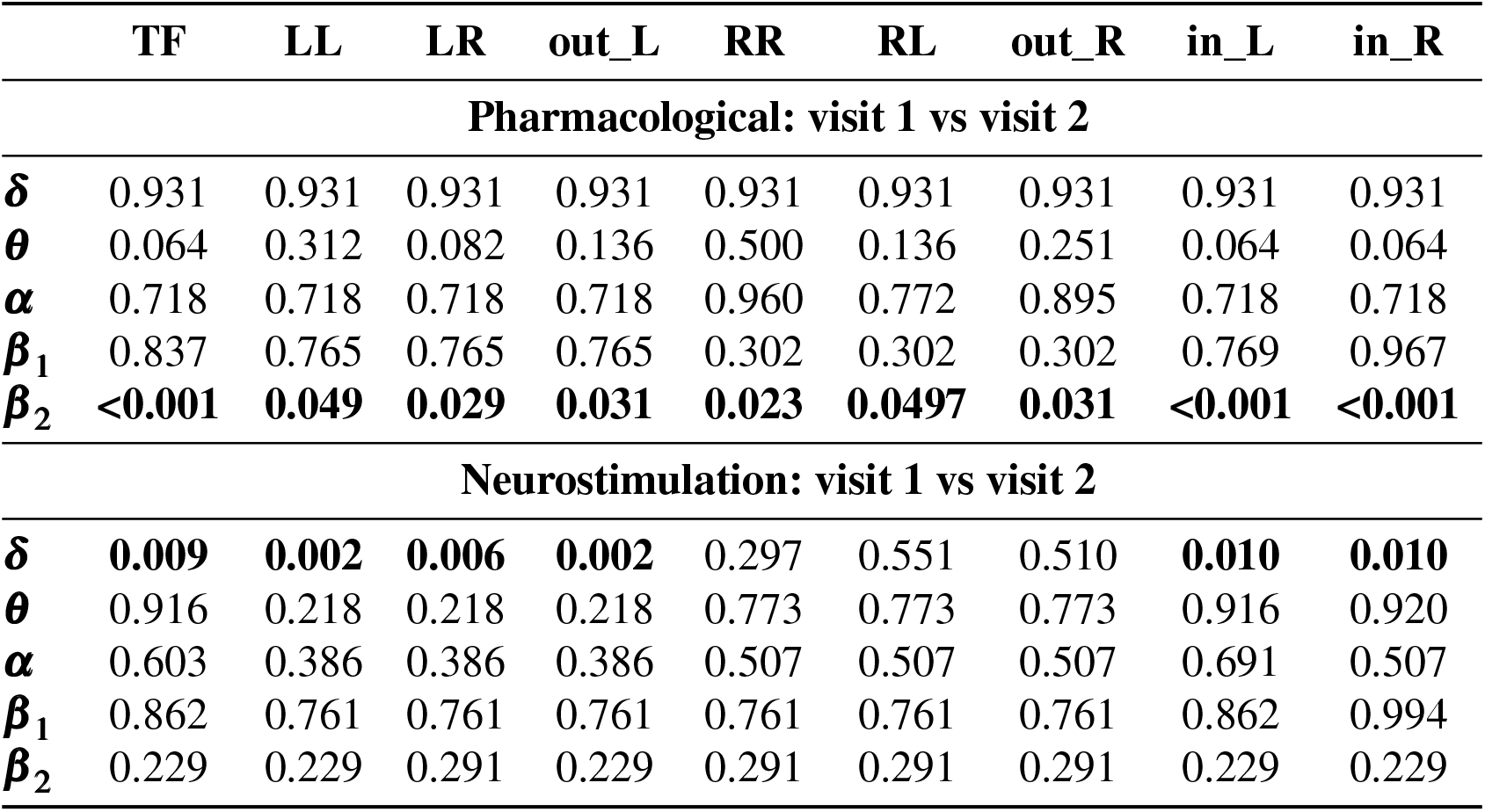
p-values for the comparison of the FLOW metrics between visits 1 and 2 for the two treatment groups. p-values *<* 0.05 are in bold font.

**Fig. 4.**
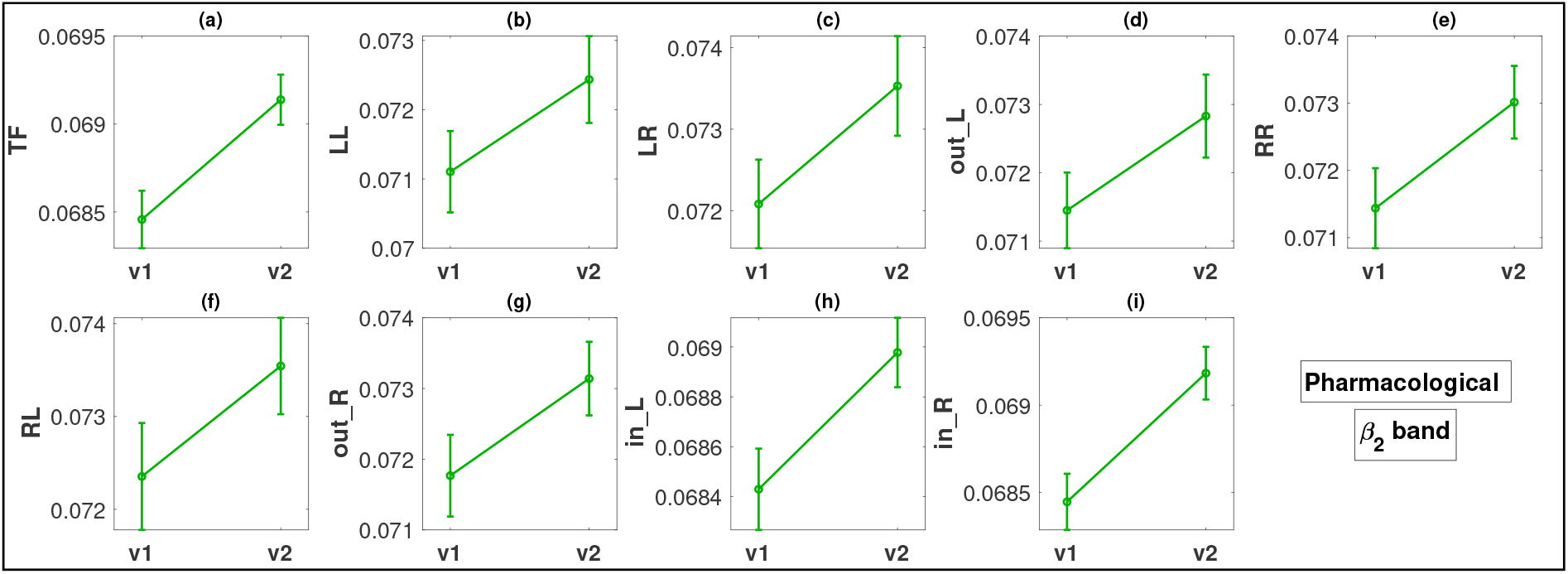
Change in metric values between visit 1 (v1) and visit 2 (v2) in the *β*_2_ band for the pharmacological treatment. The y-axis of all panels displays the metric values averaged over all subjects along with the standard errors.

**Fig. 5.**
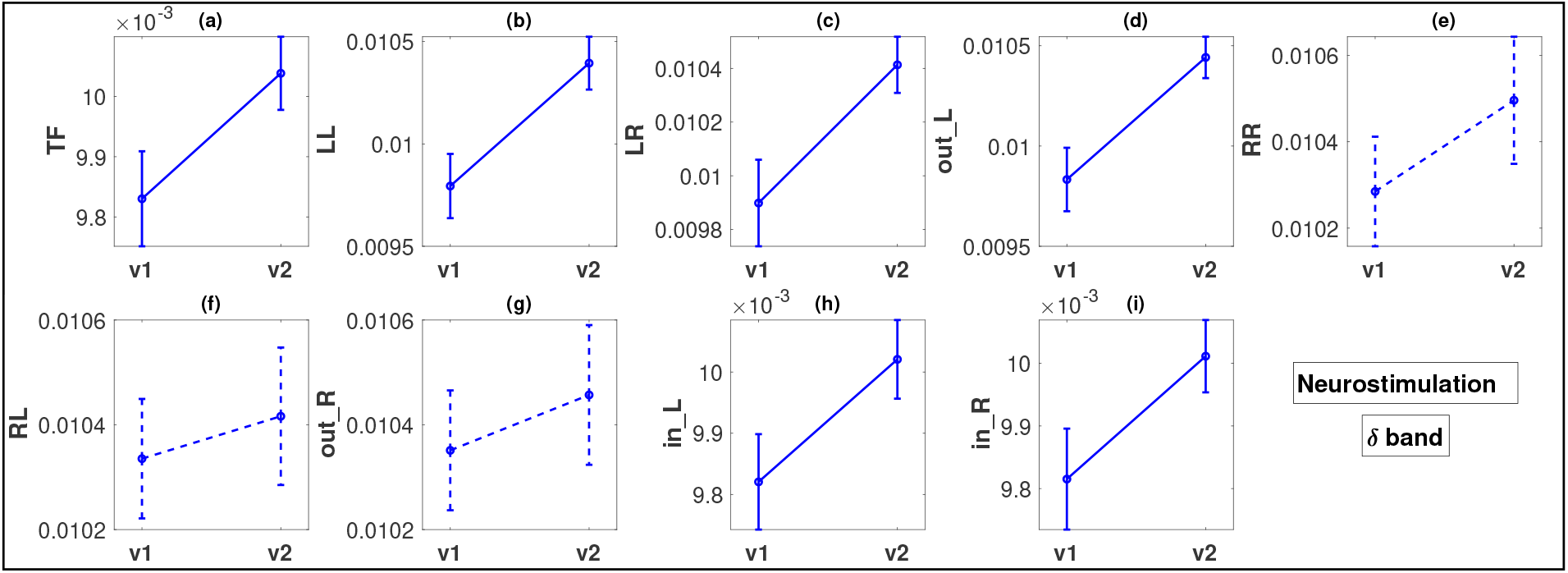
Similar to Figure 4, but for the neurostimulation treatments and the *δ* band. The dashed lines in panels (e)-(g) indicate that there was no statistically significant difference in these cases.

For the pharmacological group, in the *β*_2_ band, all the FLOW metrics (Figure 4 panels (a)- (i)) show a statistically significant increase from visit 1 to visit 2 with p-values ranging between *<* 0.001 and 0.049. The smallest p-values are observed for metrics *TF, in*_*L*, and *in*_*R*, all less than 0.001. The effect size for these three metrics based on Cohen’s *d* is moderate, or close to moderate (*d* = 0.3885, 0.3166, 0.4138, respectively). For the remaining metrics, the effect size is small (*d* in the range of 0.1900 − 0.2429).

In the case of the neurostimulation group and the *δ* band, most metrics, but not all, show significant changes. The FLOW metrics *TF, LL, LR, out*_*L, in*_*L*, and *in*_*R* exhibit a statistically significant increase between the two visits with very low p-values, at a maximum of 0.01 (Figure 5, panels (a)-(d), (h) and (i)). We note that the smallest p-values (0.002) correspond to flows originating from the left hemisphere (*LL* and *out*_*L*), for which the effect size is again moderate (*d* = 0.5993, 0.6540, respectively) but larger than the effect sizes observed in the pharmacological group. Metrics *LR, TF, in*_*L*, and *in*_*R* also exhibit a moderate effect size change (*d* in the range of 0.4015 − 0.5434). Contrary to what we observe for the left hemisphere, the metrics corresponding to outflow from the right hemisphere (*RR, RL* and *out*_*R*) do not exhibit any statistically significant changes (Figure 5, panels (e)-(g)).

Figures 6 and 7 illustrate the results of the change in inflow per channel between the two visits. The results are presented graphically in brain topographical maps and are given numerically in the supplementary materials, Table 5. In both figures, the first row of panels shows the p-values for the 19 channels in all five bands. The channels that show statistically significant changes in inflow from visit 1 to visit 2 are highlighted in green text. The results for the pharmacological group are presented in Figure 6. The second and third rows show topographical maps of the actual values of information flow per channel averaged over all subjects in the five frequency bands for visits 1 and 2, respectively. The fourth row shows the difference in the inflow as visit 2 minus visit 1. The *θ* and the *β*_2_ bands are the only ones in which the channels had a statistically significant change in inflow. The overall inflow per channel increases from visit 1 to visit 2 in both bands. The channels that are significant for both *β*_2_ and *θ* bands correspond to the right frontal (F4), central (Cz, Pz), left parietal (P3), posterior temporal (T5, T6), and occipital (O1, O2) regions. Fp2 and P4 are significant only in the *β*_2_ band but still have p-values quite low for the *θ* band (*p* = 0.058 both, marginally above 0.05). Comparing the topographic maps of the two visits for both the bands, it can be observed that the *β*_2_ band shows a larger increase in the inflow compared to the *θ* band. The effect size of the inflow metrics for the significant channels is around moderate (*d* in the range of 0.3184 − 0.5599) for the *β*_2_ band and small for the *θ* band (*d* in the range of 0.1262 − 0.1872). With respect to localisation and the change in inflow, for the *β*_2_ we observe that the statistically significant changes are quite widespread, excluding mainly left prefrontal and mid-temporal regions of the brain. The largest changes (smallest p-values) are observed for the midline frontal (Fz, *d* = 0.4361), parietal (P3, P4, *d* = 0.3871, 0.3824), occipital (O1, O2, *d* = 0.4025, 0.3743), and right posterior temporal (T6, *d* = 0.5599) regions. Comparing the left and right hemisphere p-values for the channels with statistically significant change (F3, C3, P3, O1, F7, T5 vs F4, C4, P4, O2, F8, T6), we see that the average p-value in the left is 0.012, while in the right it is 0.002 (almost an order of magnitude difference), indicating that there is more significant change (increase) of inflow in the right hemisphere.

**Fig. 6.**
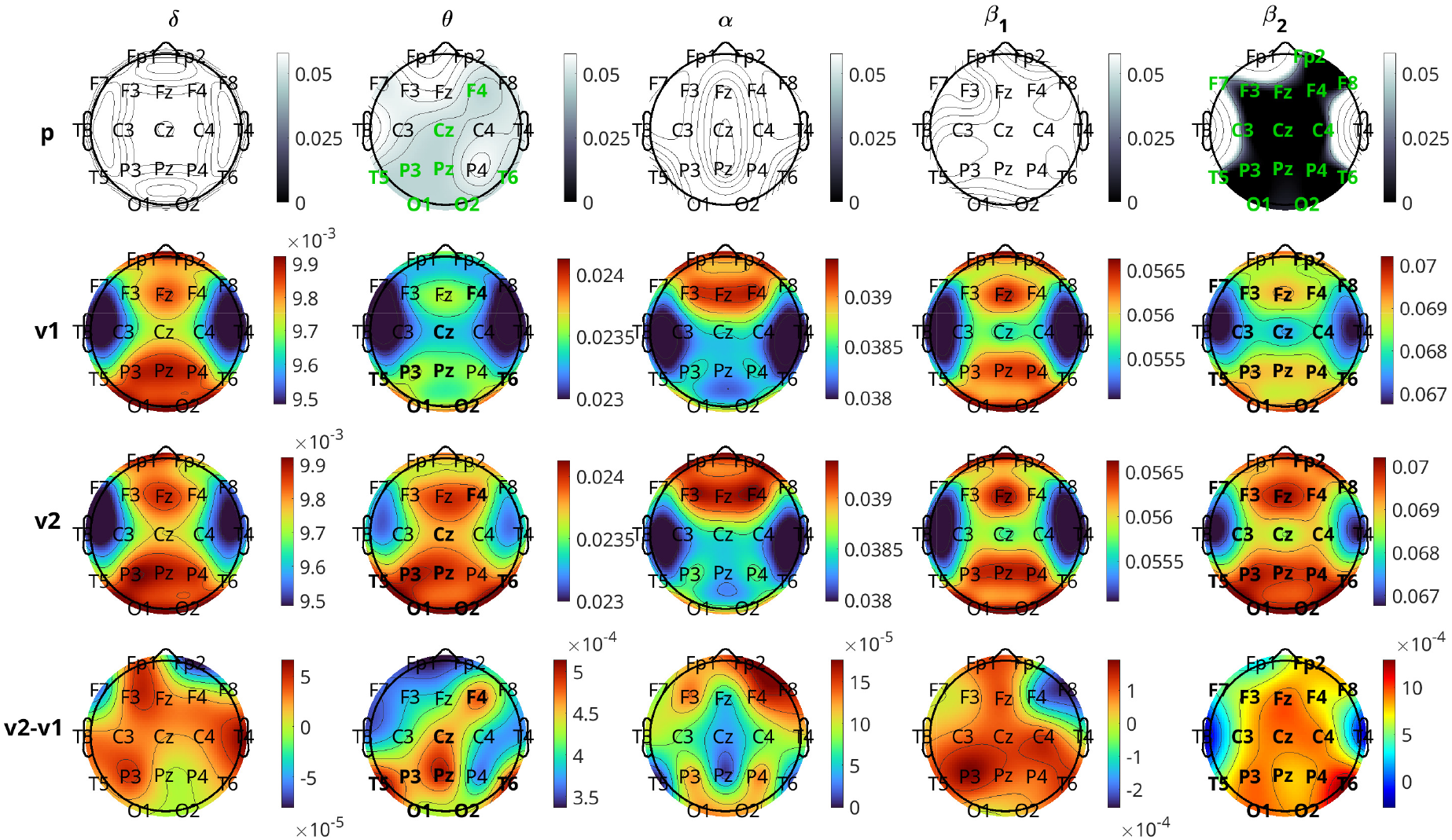
Brain topographical plots for the comparison of inflow per channel between the two visits for the pharmacological group. The first row of panels is the p-values for the comparison. The 2nd and 3rd rows present the average values (across subjects) of the inflow for visit 1 (v1) and visit 2 (v2), while the 4th row presents the difference between the visits. Columns correspond to frequency bands, as indicated by the text above the top panels. Channels with p-values *<* 0.05 are indicated in green-coloured text in the first row.

**Fig. 7.**
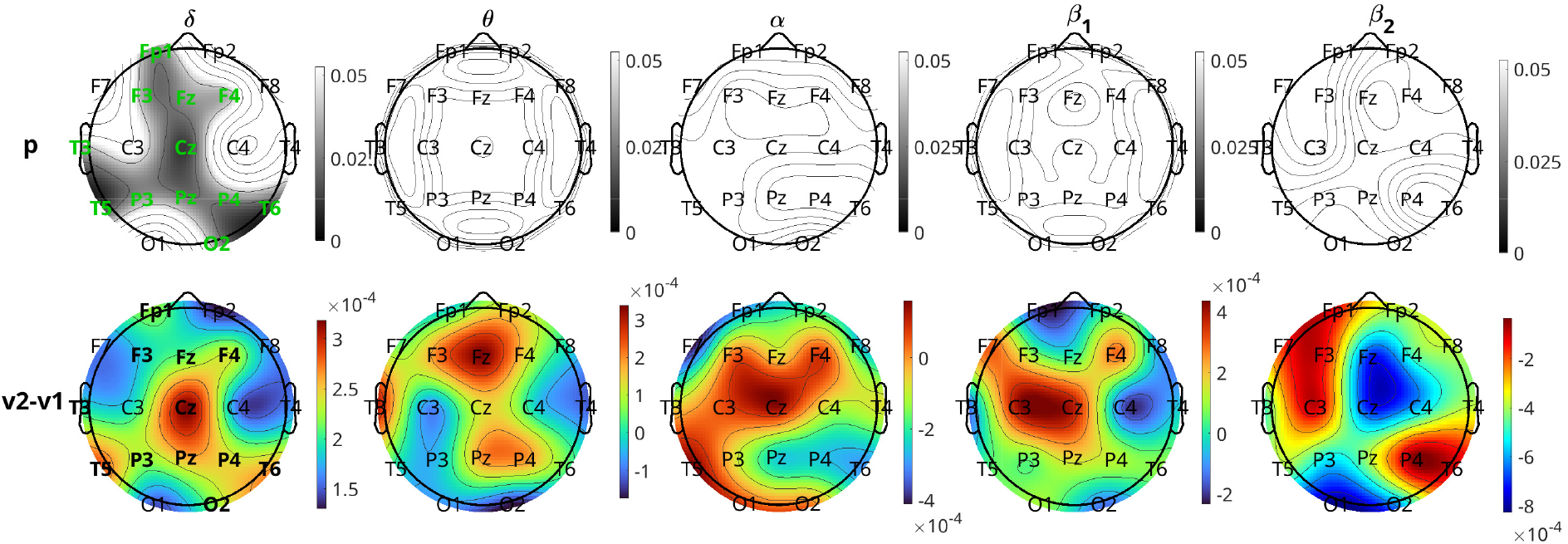
Similar to Figure 6, but for the neurostimulation group. Rows of the average inflow per channel for the two visits (2nd and 3rd rows in Figure 6) are omitted (given in supplementary materials, Figure 7).

Figure 7 presents the results for the neurostimulation group. In this case, the p-values are displayed in the first row of the panels and the difference in the average INLFOW per channel in the second row of panels. The *δ* band is the only band where a significant change in inflow is observed, with p-values of significant channels ranging between 0.013 and 0.040 (Cohen’s *d* in the range 0.2905 − 0.5348). The positive values of the difference in inflow for the *δ* band show that there is a significant increase from visit 1 to visit 2. For the *δ* band,, including the frontal (Fp1, F3, F4), central (Fz, Cz, Pz), parietal (P3, P4), occipital(O2), and temporal (T3, T5, T6) regions, but the most significant changes, i.e., the lowest p-values, are observed in the central and posterior temporal regions (Cz, T5, and T6 with *p* = 0.013 for all three and *d* = 0.5348, 0.5048, and 0.5007, respectively). Similar to pharmacological treatment, we observed that the average increase in inflow in the significant channels is greater in the right hemisphere (average *d* = 0.4205) than the left (average *d* = 0.3769).

#### 3.1.2 Baseline brain causality and response to treatment

This section presents the results of the metric analysis with respect to the response to treatment, i.e., the comparison of respondents with nonrespondents. We perform Student’s t-test (independent) to compare the metric values of visit 1 for the two response groups. The p-values for all FLOW metrics and for all five bands are listed in Table 4. Any significant difference in the metrics between the response groups will inform us about a difference in brain connectivity patterns of individuals who are going to potentially respond or not to the treatment. From Table 4, we observe that only the *α* band in the pharmacological group shows significant differences for the FLOW metrics *TF, RR, RL, out*_*R, in*_*L*, and *in*_*R* with p-values = 0.04 for all six metrics (the equality in the p-values is due to the FDR adjustment). For the neurostimulation group, there is no metric that achieves statistical significance. Prior to p-value adjustment there were three metrics in the *δ* band that showed significant change (*TF, in*_*L*, and *in*_*R* with *p* = 0.037, 0.026, and 0.041, respectively), but they did not survive the FDR procedure. The difference of the significant metric values for the pharmacological group in the *α* band is depicted in Figure 8. The y-axis of the figure displays the average over all subjects of the metrics for each response group along with their standard errors. The bar plots indicate that the nonrespondents have higher values for all these metrics than the respondents. The effect size for all these metrics is again at or near the moderate level (*d* in the range of 0.3933 − 0.4215). The bar plots corresponding to all metrics and bands are presented in the supplementary materials (Figures 8-12).

**Table 4.**
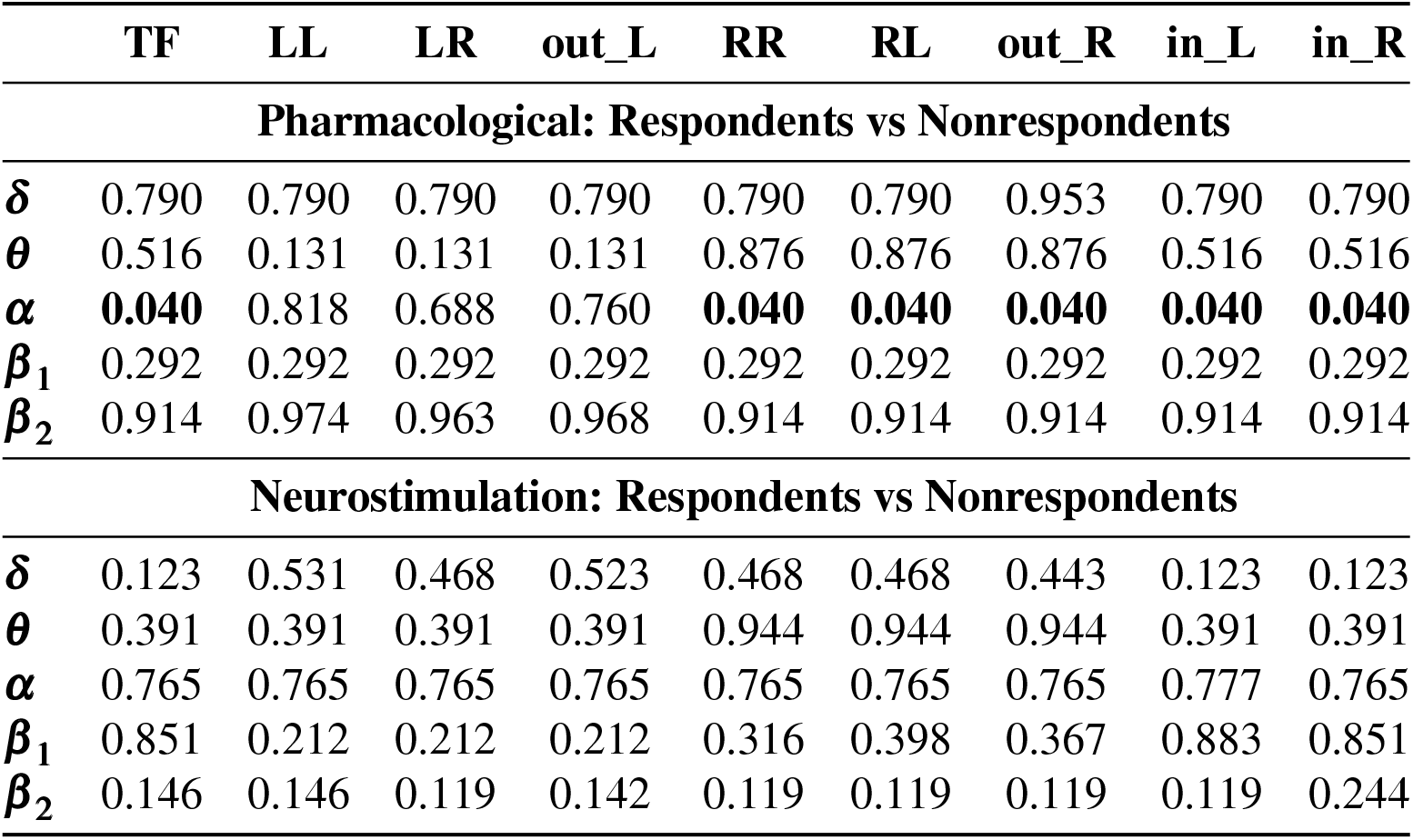
p-values for the comparison of the FLOW metrics at visit 1 between respondents and nonrespondents for the two treatment groups. Significant p-values less than 0.05 are in bold font.

**Fig. 8.**
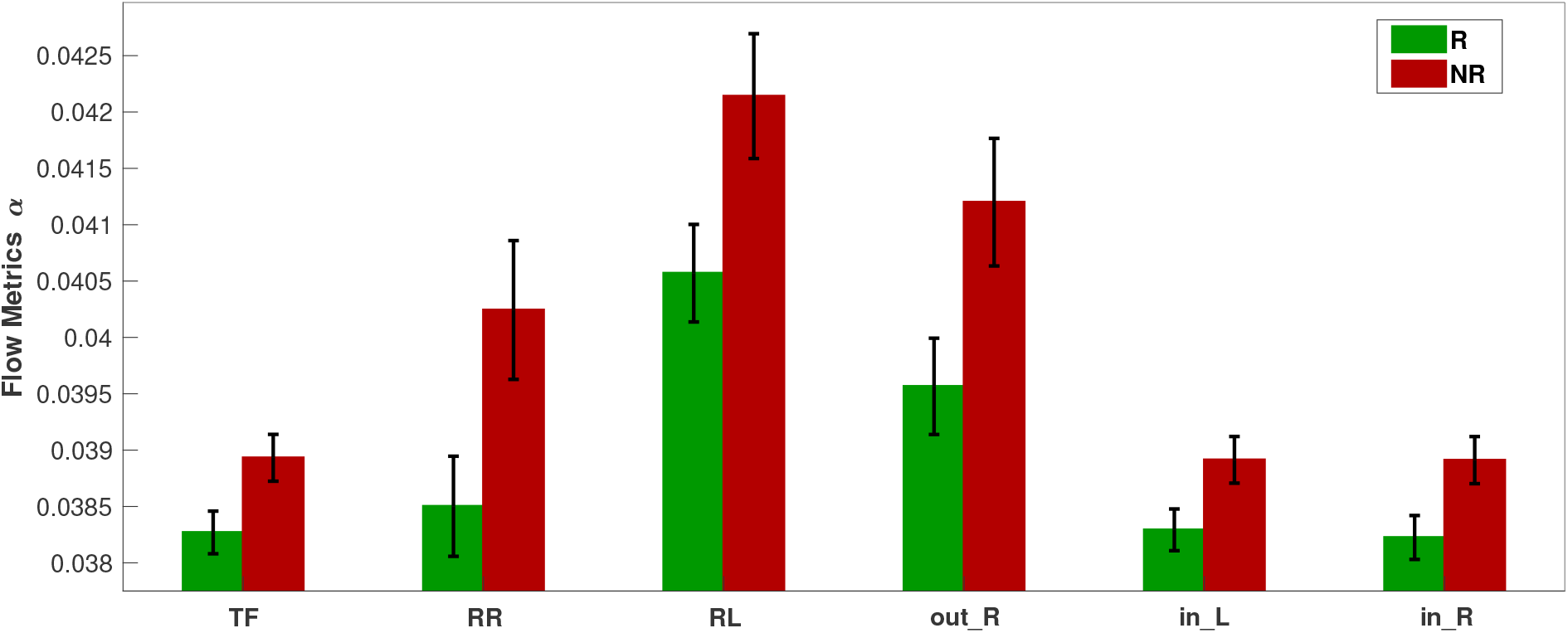
Bar plots of the significantly different FLOW metrics between respondents (R) and nonrespondents (NR) to pharmacological treatment at the time of visit 1 in the *α* band.

Figure 9 depicts the results of the comparison of inflow per channel between respondents and nonrespondents at visit 1 in all five frequency bands. The topographic plots depict the p-values for the comparisons, along with the difference in inflow averaged over all subjects. The first two rows correspond to the pharmacological group, and the last two correspond to the neurostimulation group. The *α* band is the most significant band for the pharmacological group. The most significant differences between the two response groups can be localised to the frontal (Fp1, F4), left anterior temporal (F7), right parietal (P4), right occipital (O2), and central (Pz) regions (all six having p-values = 0.048 and Cohen’s *d* = 0.431 to 0.532, indicating moderate effect size). The channels Fp2, Fz, F8, C3, C4, and P3 had p-values = 0.055, which are marginally above the significance threshold. With respect to the nature of the difference, for all the significant channels, the nonrespondents have a higher inflow per channel than the respondents. The *β*_1_ band also had two channels (Fp1 and T5) with *p <* 0.05 before FDR adjustment, but they lost significance after adjustment, and the other three bands had no significant channels. For the comparison of the response groups in the case of neurostimulation treatment, the results are not as positive. No bands show any statistically significant difference between the response groups for any channel. Prior to the FDR procedure, channels Fp1, T3, T5, C4, and O2 in the *δ* band had *p <* 0.05, but after adjustment, all of them lost significance with the adjusted p-values of C4 to be 0.082 and the rest higher. The *β*_2_ band also had three channels (F7, T3, and F4), with p-values *<* 0.05 before adjustment, which lost significance afterwards. In both bands, contrary to the pharmacological group, the respondents have a (non-significantly) higher inflow per channel compared to the nonrespondents.

**Fig. 9.**
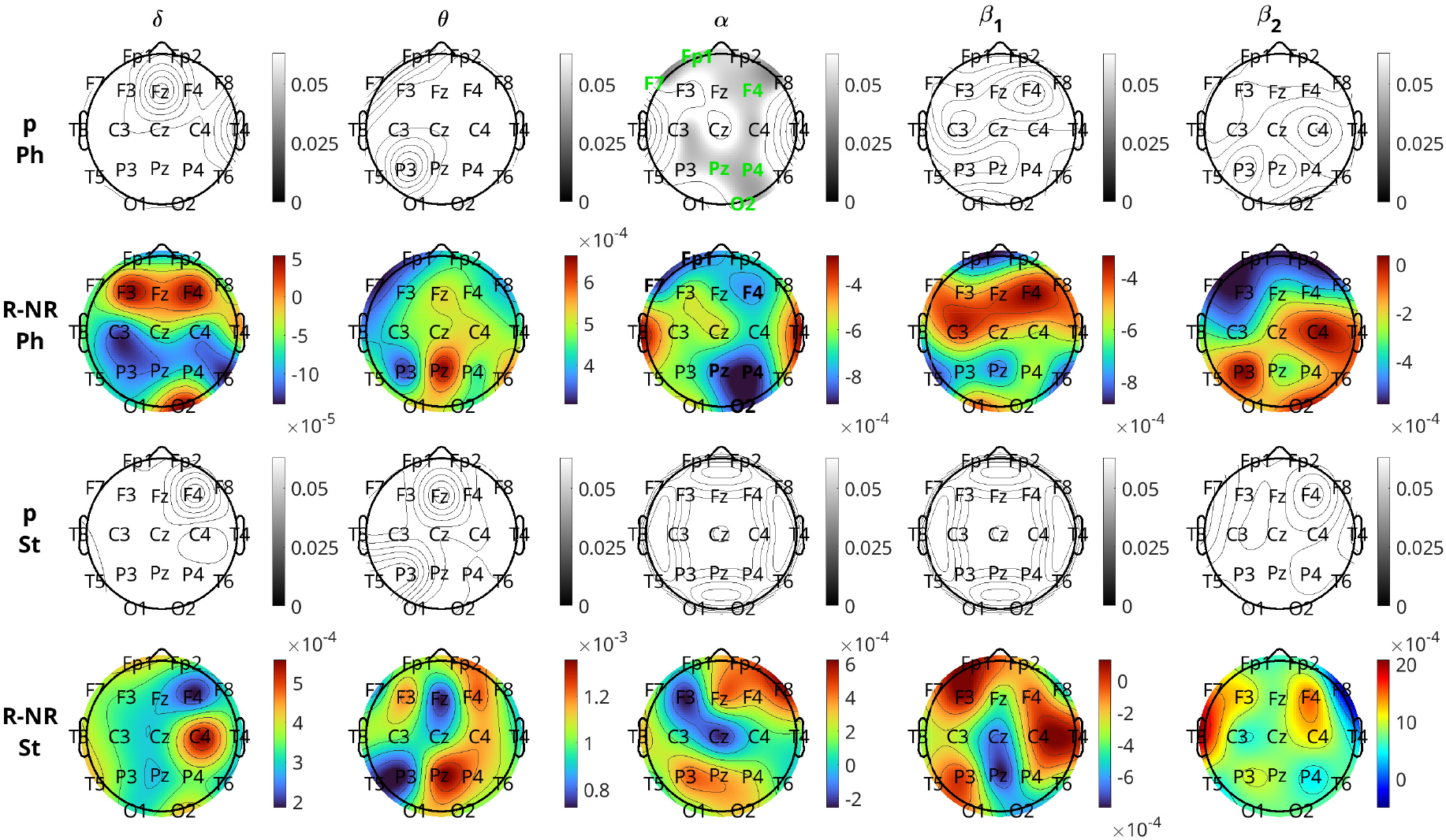
Brain topographical plots for the comparison of inflow per channel between respondents (R) and nonrespondents (NR). The first row of panels is the p-values for the pharmacological (Ph) group, and the second row is the difference in inflow between R and NR averaged over all subjects. The 3rd and 4th rows are the same as the first two but for the neurostimulation (St) group. Columns correspond to frequency bands, as indicated by the text above the top panels. Channels with p-values *<* 0.05 are indicated in green-coloured text in the 1st and 3rd rows.

## 4 Discussion

In this study, we conducted an information theory-based causality analysis on EEG data of patients diagnosed with MDD who received pharmacological or neurostimulation treatments. We introduced a slightly adjusted version of a previously developed method (pPMIME) and first tested it on artificial data, where it performed very well in capturing causal relationships for weak coupling. We then used pPMIME to estimate the pairwise strength of the causal relationships between brain locations. To study information flow in the brain, we defined sets of global (FLOW) and local (INFLOW and OUTFLOW per channel) causality metrics. We used repeated measures ANOVA to determine which factors (visit, treatment, response, and all their possible interactions) were the most important. The ANOVA results indicated overall differences between the two treatment groups and that there was no interaction between the response and visit factors. Therefore, we decided to analyse separately the data from the two treatments. We focused on two independent studies: 1) analysing the impact of each treatment modality on brain causality between visits and 2) comparing visit 1 metrics for the two response groups. The FLOW metrics performed well in differentiating the two visits as well as the two response groups, the latter only for the pharmacological treatment. With respect to the local metrics, the OUTFLOW per channel performed much worse than INFLOW. OUTFLOW provided fewer in number and magnitude significant p-values that did not provide any strong localisation information. On the other hand, INFLOW provided insight about brain region-specific information flow and indicated brain regions whose “causal behaviour” is altered by treatment and may potentially serve as indicators of response to treatment.

The first analysis of the effect of treatment on brain connectivity revealed that the pharmacological and the neurostimulation treatments lead to different types of changes in information flow in the brain. The main observation for the effect of one week of pharmacological treatment was an increase in *β*_2_ information flow in the brain. This effect is relatively global with no strong hemispheric differentiation. With respect to inflow at specific brain regions, the major changes were seen again in *β*_2_ and were maximal at the midline frontal, parietal, and occipital regions. These results may be related to the increase in *β* band power for pharmacological treatment reported in [46] and to existing findings that state that MDD is characterised by unique and dominant *β* oscillations [47]. Also, MDD is known to affect distributed large-scale cortical and subcortical systems, which are functionally interconnected across the frontal, temporal, parietal, and occipital lobes [48]. Our results also indicate that the most significant changes are seen in these regions. Additional observations regarding some left-right asymmetry in the *β*_2_ band are that the information flow from left to right is greater than that in the opposite direction. We also observed that in the *β*_2_ band, the inflow to certain regions of the right hemisphere increased more compared to the left hemisphere at visit 2. This is in agreement with the belief that the right hemisphere is affected more in MDD [49, 50, 51, 52].

For the neurostimulation group, the *δ* band captured the difference between the two visits as a statistically significant increase in global information flow from the left hemisphere to the whole brain. This may be related to results from certain studies where changes in brain connectivity were reported in the *δ* band due to rTMS [53, 54]. With respect to the inflow at specific brain regions, the changes were widespread, with the central and posterior temporal regions showing the larger difference. Previous studies [53, 54] that concentrated only on the frontal lobe channels of MDD patients have reported significant changes due to the treatment. Our results also showed significant differences in the frontal lobe channels (Fp1, F3, Fz, and F4).

The visit 1 data analysis revealed that the potential responders and nonresponders to pharmacological treatment differed most in the *α* band. This result relates to research that has reported *α* band differences between responders and nonresponders to therapy using activity and connectivity measures [55] and to *α* band functional connectivity increase in MDD [56]. Our results show that for this treatment group, the nonrespondents have higher *α* band information flow from right to left and a higher total inflow to each hemisphere. Previous studies from pre-treatment EEG data have reported that nonresponders to pharmacological treatment of type SSRI showed greater activation of the right hemisphere than the left in the *α* band. [57, 58]. At the local level, the *α* band captured most differences in inflow per channel between the response groups. While the significant channels were widespread, the frontal, right parietal, left anterior temporal and right occipital regions showed the largest difference, with the nonrespondents showing higher inflow. These results could be related to earlier research, which reported higher *α* band activity in patients with depression in large portions of the brain [57, 55].

For the neurostimulation group, as discussed earlier, the results were not very positive; however, there were some interesting observations. Three FLOW metrics in the *δ* band, corresponding to the total flow (*TF*), inflow to the left (*in*_*L*), and the right hemisphere (*in*_*R*), showed the biggest difference between the potential respondents and the nonrespondents. For all three metrics, the respondents had higher metric values than the nonrespondents (see supplementary materials, Figure 10). This is in accordance with the work by Nobakhsh et al. [53], which identified effective brain connectivity in the *δ* band as a valuable biomarker for distinguishing respondents and nonrespondents in drug-resistant MDD patients, based on baseline data, following rTMS treatment. At the local level, the respondents had a higher inflow per channel. The study by Nobakhsh et al. [53] also reported that respondents had higher activity in the prefrontal cortex. In our case, unfortunately, the neurostimulation group had a small sample size (*n* = 47, with 16 respondents and 31 nonrespondents), which decreases statistical power. As a matter of fact, Cohen’s *d* for the three above-mentioned metrics were high (0.6501, 0.6986, and 0.6364, respectively) and, in fact, higher than the ones observed in the pharmacological group. Similar results were observed at the local level, where respondents had a higher inflow per channel than the nonrespondents with *d* in the range 0.643 − 0.910 (moderate to large effect size) for the channels that had *p <* 0.05 before FDR adjustment. We expect that if a larger sample were available, these metrics would be deemed significant even after p-value adjustment.

The findings of our study complement the existing results in the literature, which has established that several brain regions and frequency bands are affected in MDD [56, 59, 60, 61]. Certain brain regions in the right hemisphere are known to be affected more in MDD, and research involving neuroimaging [50, 51] and electrophysiological measures [52, 49] has shown that asymmetric activation between the right and left brain regions is significant. We have also observed that the effect size of the information flow metrics to the right hemisphere is larger than the left between visits for both treatment methods. However, a limitation of our work is that these observations related to the hemispheric asymmetry are not conclusive, as they are not based on statistical testing. We plan to explore this as future work, where we will concentrate explicitly on global and local asymmetry metrics. Another limitation is that we had comparatively small sample sizes for the neurostimulation treatment, which probably led to difficulty in the statistical comparisons for this treatment group.

## 5 Conclusions

Our work aligns with existing research and provides novel results in the direction of causal influence in the brain in MDD. The study’s findings show that the two treatment modalities affected brain information flow differently. We observed a significant imbalance in brain connection patterns and information flow between individuals likely to respond to treatment and those unlikely to respond. This imbalance differs between the two treatment modalities. This study demonstrated synergy with existing literature by showing that multiple brain regions are affected in MDD. The information flow metrics central to our study can serve as valuable biomarkers that can augment the development of effective therapeutic approaches.

## Data Availability

All data produced in the present study are available upon reasonable request to the authors

## Data availability statement

The data used in this study are available upon reasonable request.

## Ethics statement

This study was conducted in accordance with the principles outlined in the Declaration of Helsinki and adhered to applicable guidelines for Good Clinical Practice. Ethical approval was obtained from the Ethics Committee of the Prague Psychiatric Centre/National Institute of Mental Health for all study protocols contributing data to this research. While a portion of the dataset was derived from clinical trials registered in the EUDRACT database (EUDRACT Nos. 2005- 000826-22 and 2015-001639-19, registered via www.clinicaltrialsregister.eu), additional data were obtained from ethically approved studies that were not formally registered in public trial registries. In all cases, adult participants were thoroughly informed about the study procedures and objectives, and written informed consent was obtained prior to participation.

## Acknowledgments

This study is supported by the Czech Science Foundation, Project No. GF21-14727K, and the Czech Academy of Sciences, Praemium Academiae awarded to M. Paluš. It was also supported by ERDF-Project Brain dynamics, No. CZ.02.01.01/00/22_008/0004643 and the Czech Science Foundation project No. 23-07074S awarded to J. Hlinka and Charles University Program Cooperatio 38:Neurosciences awarded to Martin Brunovský.

