## supplementary materials for "Causality Analysis in Major Depressive Disorder for Early Prediction of Treatment Outcomes with Pharmacological and Neuromodulation Therapies"

Madhurima Bhattacharjee[1], Ioannis Vlachos[1,2,3], Aditi Kathpalia*[1], Jaroslav Hlinka[1], Martin Brunovský[4,5], Martin Bareš[4,5], and Milan Paluš[1]

[1]Department of Complex Systems, Institute of Computer Science of the Czech Academy of Sciences, Prague,Czech Republic
[2]Department of Electrical and Computer Engineering, Aristotle University of Thessaloniki, 54124 Thessaloniki, Greece
[3]Medical School, Aristotle University of Thessaloniki, Thessaloniki 54124, Greece
[4]Clinical Research Programme, National Institute of Mental Health, Klecany, Czech Republic
[5]Charles University, Third Faculty of Medicine, Prague, Czech Republic

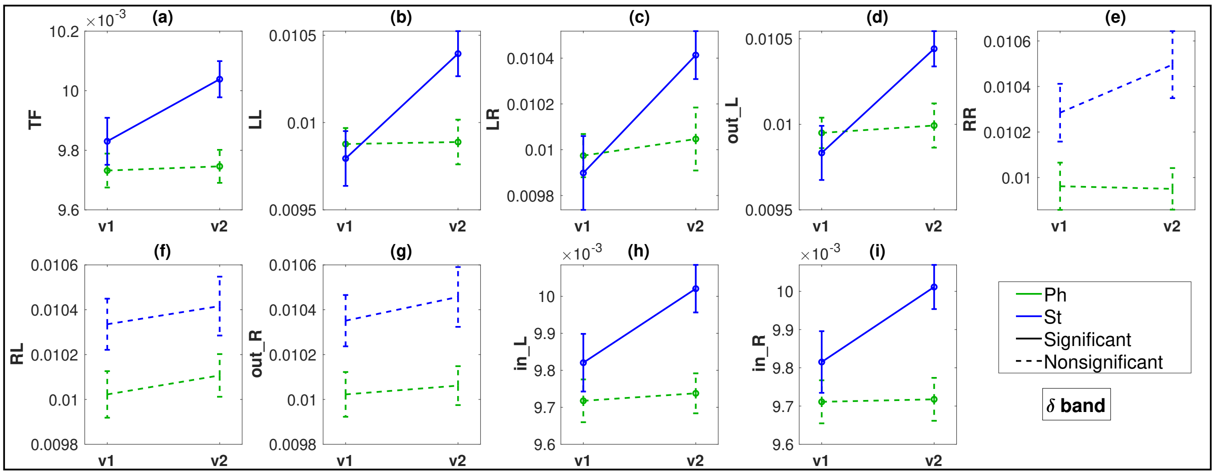


Figure 1: $\delta$ band: FLOW Metric comparisons between visits 1 (v1) and 2 (v2) for the pharmacological (Ph) and neurostimulation (St) groups. Metrics with $p < 0.05$ plotted in bold lines and $p \geq 0.05$ are in dotted lines.

*Department of Applied Mechanics and Biomedical Engineering, Indian Institute of Technology, Madras, Chennai, India

Table 1: Repeated measure ANOVA FLOW metric results. Significant p values less than 0.05 are in bold font.

| | **TF** | **LL** | **LR** | **out_L** | **RR** | **RL** | **out_R** | **in_L** | **in_R** |
|---|---|---|---|---|---|---|---|---|---|
| | | | | | **RESPONSE** | | | | |
| $\delta$ | 0.183 | 0.398 | 0.656 | 0.560 | 0.230 | 0.667 | 0.304 | 0.152 | 0.212 |
| $\theta$ | 0.127 | **0.047** | 0.083 | 0.062 | 0.889 | 0.822 | 0.805 | 0.134 | 0.125 |
| $\alpha$ | 0.155 | 0.796 | 0.634 | 0.674 | 0.127 | 0.082 | 0.106 | 0.143 | 0.207 |
| $\beta_1$ | 0.113 | 0.103 | 0.055 | 0.072 | 0.627 | 0.878 | 0.771 | 0.137 | 0.113 |
| $\beta_2$ | 0.167 | 0.094 | 0.114 | 0.106 | **0.021** | **0.048** | **0.033** | 0.124 | 0.182 |
| | | | | | **TREATMENT** | | | | |
| $\delta$ | **0.012** | 0.186 | 0.372 | 0.290 | **0.002** | **0.032** | **0.007** | **0.011** | **0.011** |
| $\theta$ | 0.123 | 0.317 | 0.156 | 0.198 | 0.401 | 0.267 | 0.319 | 0.123 | 0.120 |
| $\alpha$ | 0.444 | **0.013** | 0.064 | **0.032** | 0.955 | 0.757 | 0.812 | 0.407 | 0.456 |
| $\beta_1$ | 0.912 | 0.067 | 0.116 | 0.088 | 0.249 | 0.173 | 0.205 | 0.855 | 0.855 |
| $\beta_2$ | 0.234 | **0.018** | 0.055 | **0.031** | 0.550 | 0.381 | 0.468 | 0.272 | 0.254 |
| | | | | | **VISIT** | | | | |
| $\delta$ | 0.099 | **0.028** | 0.061 | **0.020** | 0.346 | 0.510 | 0.523 | 0.108 | 0.155 |
| $\theta$ | 0.155 | 0.066 | **0.025** | **0.032** | 0.829 | 0.667 | 0.853 | 0.160 | 0.226 |
| $\alpha$ | 0.713 | 0.253 | 0.269 | 0.284 | 0.478 | 0.595 | 0.540 | 0.835 | 0.649 |
| $\beta_1$ | 0.976 | 0.402 | 0.326 | 0.340 | 0.750 | 0.913 | 0.943 | 0.929 | 0.650 |
| $\beta_2$ | 0.191 | 0.948 | 0.645 | 0.924 | **0.032** | 0.065 | **0.044** | 0.370 | 0.082 |
| | | | | | **RESPONSE:TREATMENT** | | | | |
| $\delta$ | 0.263 | 0.626 | 0.972 | 0.815 | 0.581 | 0.424 | 0.411 | 0.216 | 0.278 |
| $\theta$ | 0.479 | 0.721 | 0.720 | 0.662 | 0.965 | 0.929 | 0.961 | 0.466 | 0.479 |
| $\alpha$ | 0.288 | 0.361 | 0.323 | 0.362 | 0.757 | 0.752 | 0.713 | 0.333 | 0.259 |
| $\beta_1$ | 0.918 | **0.007** | **0.004** | **0.005** | 0.074 | 0.101 | 0.081 | 0.948 | 0.889 |
| $\beta_2$ | **0.038** | 0.181 | 0.070 | 0.116 | **0.011** | **0.020** | **0.017** | **0.007** | 0.084 |
| | | | | | **RESPONSE:VISIT** | | | | |
| $\delta$ | 0.669 | 0.970 | 0.973 | 0.890 | 0.916 | 0.722 | 0.983 | 0.739 | 0.512 |
| $\theta$ | 0.672 | 0.527 | 0.219 | 0.329 | 0.970 | 0.948 | 0.945 | 0.862 | 0.536 |
| $\alpha$ | 0.196 | 0.077 | **0.040** | 0.056 | 0.508 | 0.960 | 0.808 | 0.196 | 0.269 |
| $\beta_1$ | 0.267 | 0.809 | 0.873 | 0.837 | 0.584 | 0.712 | 0.710 | 0.205 | 0.256 |
| $\beta_2$ | 0.304 | 0.938 | 0.521 | 0.801 | 0.745 | 0.613 | 0.623 | 0.478 | 0.137 |
| | | | | | **TREATMENT:VISIT** | | | | |
| $\delta$ | 0.143 | **0.030** | 0.153 | **0.038** | 0.307 | 0.936 | 0.781 | 0.185 | 0.172 |
| $\theta$ | 0.260 | 0.514 | 0.982 | 0.754 | 0.366 | 0.177 | 0.254 | 0.270 | 0.196 |
| $\alpha$ | 0.255 | 0.794 | 0.955 | 0.911 | 0.554 | 0.305 | 0.409 | 0.322 | 0.171 |
| $\beta_1$ | 0.734 | 0.967 | 0.784 | 0.800 | **0.050** | 0.100 | 0.070 | 0.668 | 0.697 |
| $\beta_2$ | **0.0001** | **0.031** | **0.047** | **0.028** | 0.657 | 0.904 | 0.773 | **0.003** | **0.0001** |
| | | | | | **RESPONSE:TREATMENT:VISIT** | | | | |
| $\delta$ | 0.079 | 0.222 | 0.050 | 0.099 | 0.745 | 0.333 | 0.691 | 0.069 | 0.085 |
| $\theta$ | 0.874 | 0.637 | 0.994 | 0.839 | 0.916 | 0.821 | 0.853 | 0.801 | 0.983 |
| $\alpha$ | 0.283 | 0.781 | 0.490 | 0.649 | 0.575 | 0.492 | 0.501 | 0.387 | 0.208 |
| $\beta_1$ | 0.134 | 0.997 | 0.846 | 0.880 | 0.368 | 0.545 | 0.461 | 0.103 | 0.124 |
| $\beta_2$ | 0.716 | 0.488 | 0.405 | 0.512 | 0.793 | 0.912 | 0.890 | 0.586 | 0.563 |

Table 2: Repeated measure ANOVA INFLOW per channel metric results. Significant p values less than 0.05 are in bold font.

| | Fp1 | F3 | C3 | P3 | O1 | F7 | T3 | T5 | Fp2 | F4 | C4 | P4 | O2 | F8 | T4 | T6 | Fz | Cz | Pz |
|---|---|---|---|---|---|---|---|---|---|---|---|---|---|---|---|---|---|---|---|
| RESPONSE | | | | | | | | | | | | | | | | | | | |
| $\delta$ | 0.153 | 0.208 | 0.595 | 0.164 | 0.320 | 0.126 | 0.093 | 0.105 | 0.474 | 0.371 | 0.158 | 0.136 | 0.067 | 0.198 | 0.244 | 0.730 | 0.408 | 0.104 | 0.321 |
| $\theta$ | 0.133 | 0.105 | 0.125 | 0.218 | 0.155 | 0.156 | 0.089 | 0.156 | 0.137 | 0.103 | 0.101 | 0.086 | 0.259 | 0.126 | 0.139 | 0.129 | 0.196 | 0.174 | **0.048** |
| $\alpha$ | 0.126 | **0.042** | 0.146 | 0.239 | 0.225 | 0.062 | 0.664 | 0.256 | 0.398 | 0.191 | 0.221 | 0.161 | 0.129 | 0.375 | 0.734 | 0.081 | 0.105 | 0.072 | 0.205 |
| $\beta_1$ | 0.216 | 0.221 | 0.173 | 0.129 | 0.122 | 0.237 | 0.077 | 0.120 | 0.116 | 0.165 | 0.207 | 0.087 | 0.116 | 0.128 | 0.203 | 0.075 | 0.168 | 0.056 | 0.054 |
| $\beta_2$ | 0.376 | 0.473 | 0.316 | 0.091 | 0.228 | **0.038** | 0.392 | 0.281 | 0.530 | 0.169 | **0.042** | 0.334 | 0.130 | 0.936 | 0.377 | 0.214 | 0.192 | 0.868 | 0.228 |
| TREATMENT | | | | | | | | | | | | | | | | | | | |
| $\delta$ | **0.002** | **0.014** | 0.140 | **0.023** | **0.010** | **0.006** | **0.011** | 0.201 | **0.001** | **0.023** | **0.027** | **0.010** | **0.023** | **0.008** | 0.064 | 0.186 | **0.030** | **0.026** | **0.046** |
| $\theta$ | 0.096 | 0.064 | 0.132 | 0.222 | 0.189 | 0.101 | 0.073 | 0.222 | 0.140 | 0.116 | 0.091 | 0.106 | 0.221 | 0.099 | 0.091 | 0.165 | 0.151 | 0.131 | 0.134 |
| $\alpha$ | 0.452 | 0.392 | 0.646 | 0.438 | 0.339 | 0.571 | 0.414 | 0.338 | 0.993 | 0.551 | 0.375 | 0.435 | 0.328 | 0.776 | 0.254 | 0.335 | 0.877 | 0.553 | 0.330 |
| $\beta_1$ | 0.823 | 0.727 | 0.915 | 0.995 | 0.857 | 0.854 | 0.754 | 0.340 | 0.837 | 0.660 | 0.581 | 0.715 | 0.928 | 0.807 | 0.532 | 0.746 | 0.987 | 0.896 | 0.398 |
| $\beta_2$ | 0.302 | **0.032** | 0.171 | 0.158 | 0.197 | 0.158 | 0.493 | 0.142 | 0.450 | 0.119 | 0.138 | 0.495 | 0.087 | 0.112 | 0.130 | 0.236 | 0.082 | 0.393 | 0.299 |
| VISIT | | | | | | | | | | | | | | | | | | | |
| $\delta$ | 0.135 | 0.125 | 0.247 | 0.093 | 0.371 | 0.589 | 0.173 | **0.044** | 0.951 | 0.100 | 0.541 | 0.069 | 0.165 | 0.346 | 0.141 | 0.055 | 0.127 | **0.015** | 0.107 |
| $\theta$ | 0.313 | 0.112 | 0.421 | 0.185 | 0.348 | 0.169 | 0.065 | 0.102 | 0.205 | 0.133 | 0.378 | 0.093 | 0.619 | 0.229 | 0.446 | 0.176 | **0.041** | 0.086 | 0.081 |
| $\alpha$ | 0.457 | 0.674 | 0.756 | 0.778 | 0.747 | 0.131 | 0.872 | 0.777 | 0.748 | 0.740 | 0.875 | 0.504 | 0.940 | 0.491 | 0.966 | 0.134 | 0.523 | 0.634 | 0.301 |
| $\beta_1$ | 0.478 | 0.887 | 0.193 | 0.808 | 0.792 | 0.515 | 0.567 | 0.918 | 0.615 | 0.830 | 0.499 | 0.836 | 0.779 | 0.241 | 0.385 | 0.705 | 0.881 | 0.255 | 0.568 |
| $\beta_2$ | 0.427 | 0.217 | 0.181 | 0.277 | 0.581 | 0.401 | 0.125 | 0.382 | 0.228 | 0.684 | 0.526 | **0.020** | 0.226 | 0.293 | 0.447 | **0.003** | 0.290 | 0.946 | 0.269 |
| RESPONSE:TREATMENT | | | | | | | | | | | | | | | | | | | |
| $\delta$ | 0.161 | 0.411 | 0.463 | 0.319 | 0.272 | 0.232 | 0.201 | 0.132 | 0.328 | 0.984 | 0.213 | 0.110 | 0.273 | 0.209 | 0.573 | 0.247 | 0.439 | 0.487 | 0.364 |
| $\theta$ | 0.347 | 0.409 | 0.625 | 0.490 | 0.582 | 0.384 | 0.406 | 0.567 | 0.414 | 0.468 | 0.549 | 0.300 | 0.648 | 0.401 | 0.523 | 0.615 | 0.633 | 0.534 | 0.413 |
| $\alpha$ | 0.303 | 0.507 | 0.284 | 0.217 | 0.404 | 0.294 | 0.428 | 0.584 | 0.163 | 0.284 | 0.232 | 0.176 | 0.335 | 0.099 | 0.558 | 0.911 | 0.255 | 0.656 | 0.161 |
| $\beta_1$ | 0.536 | 0.908 | 0.861 | 0.897 | 0.696 | 0.913 | 0.974 | 0.873 | 0.743 | 0.793 | 0.934 | 0.997 | 0.973 | 0.690 | 0.865 | 0.996 | 0.775 | 0.633 | 0.614 |
| $\beta_2$ | **0.013** | **0.023** | 0.099 | 0.136 | 0.151 | **0.006** | **0.009** | 0.065 | 0.088 | **0.020** | 0.135 | 0.295 | 0.122 | 0.731 | 0.331 | 0.105 | 0.075 | 0.879 | 0.131 |
| RESPONSE:VISIT | | | | | | | | | | | | | | | | | | | |
| $\delta$ | 0.708 | 0.387 | 0.479 | 0.580 | 0.802 | 0.935 | 0.665 | 0.892 | 0.397 | 0.721 | 0.120 | 0.721 | 0.420 | 1.000 | 0.552 | 0.527 | 0.358 | 0.592 | 0.755 |
| $\theta$ | 0.933 | 0.633 | 0.692 | 0.944 | 0.394 | 0.909 | 0.938 | 0.812 | 0.381 | 0.585 | 0.447 | 0.761 | 0.215 | 0.895 | 0.723 | 0.631 | 0.813 | 0.592 | 0.462 |
| $\alpha$ | 0.476 | 0.196 | 0.370 | 0.095 | 0.228 | 0.247 | 0.303 | 0.525 | 0.461 | 0.271 | 0.652 | 0.517 | 0.285 | 0.111 | 0.988 | 0.071 | **0.017** | 0.418 | 0.340 |
| $\beta_1$ | 0.595 | 0.151 | 0.266 | 0.223 | 0.059 | 0.329 | 0.503 | 0.562 | 0.372 | **0.049** | 0.221 | 0.153 | 0.790 | 0.881 | 0.180 | 0.728 | 0.479 | 0.432 | 0.976 |
| $\beta_2$ | 0.199 | 0.767 | 0.321 | 0.560 | 0.282 | 0.177 | 0.088 | 0.301 | 0.365 | 0.800 | 0.634 | 0.204 | 0.266 | **0.049** | 0.313 | 0.598 | 0.133 | 0.319 | 0.351 |
| TREATMENT:VISIT | | | | | | | | | | | | | | | | | | | |
| $\delta$ | 0.351 | 0.416 | 0.403 | 0.271 | 0.392 | 0.107 | 0.315 | 0.075 | 0.287 | 0.160 | 0.827 | 0.075 | 0.146 | 0.233 | 0.634 | 0.123 | 0.288 | **0.027** | 0.089 |
| $\theta$ | 0.352 | 0.578 | 0.121 | 0.179 | 0.101 | 0.386 | 0.888 | 0.179 | 0.400 | 0.271 | 0.161 | 0.631 | **0.042** | 0.254 | 0.138 | 0.189 | 0.890 | 0.317 | 0.364 |
| $\alpha$ | 0.141 | 0.606 | 0.828 | 0.251 | 0.599 | **0.041** | 0.384 | 0.748 | 0.149 | 0.505 | 0.454 | 0.138 | 0.422 | 0.078 | 0.543 | 0.102 | 0.311 | 0.833 | 0.274 |
| $\beta_1$ | 0.187 | 0.756 | 0.468 | 0.393 | 0.754 | 0.514 | 0.398 | 0.558 | 0.607 | 0.394 | 0.172 | 0.742 | 0.554 | 0.738 | 0.606 | 0.701 | 0.613 | 0.601 | 0.974 |
| $\beta_2$ | 0.600 | **0.034** | 0.206 | **0.001** | **2.27e-05** | 0.239 | 0.474 | **0.044** | **0.003** | **0.006** | **0.004** | **0.043** | **3.24e-04** | 0.062 | 0.997 | **0.002** | **8.13E-06** | **0.001** | **0.007** |
| RESPONSE:TREATMENT:VISIT | | | | | | | | | | | | | | | | | | | |
| $\delta$ | 0.127 | 0.435 | **0.020** | 0.106 | 0.272 | 0.098 | 0.060 | 0.182 | 0.126 | 0.351 | **0.001** | 0.444 | 0.258 | 0.338 | 0.142 | 0.115 | 0.403 | 0.079 | 0.198 |
| $\theta$ | 0.659 | 0.918 | 0.695 | 0.563 | 0.906 | 0.568 | 0.908 | 0.411 | 0.918 | 0.751 | 0.701 | 0.599 | 0.628 | 0.679 | 0.812 | 0.990 | 0.547 | 0.668 | 0.957 |
| $\alpha$ | 0.310 | 0.795 | 0.869 | 0.237 | 0.528 | 0.180 | 0.536 | 0.246 | 0.371 | 0.106 | 0.874 | 0.340 | **0.048** | 0.387 | 0.797 | 0.085 | 0.162 | 0.854 | 0.372 |
| $\beta_1$ | 0.129 | 0.110 | 0.565 | 0.091 | 0.094 | 0.309 | 0.311 | 0.067 | **0.018** | 0.161 | 0.094 | 0.347 | 0.902 | 0.537 | **0.032** | 0.247 | 0.488 | 0.575 | 0.294 |
| $\beta_2$ | 0.131 | 0.700 | 0.749 | 0.693 | 0.610 | 0.818 | 0.532 | 0.116 | 0.947 | 0.627 | 0.867 | 0.770 | 0.244 | 0.563 | 0.460 | 0.501 | 0.546 | 0.122 | 0.967 |

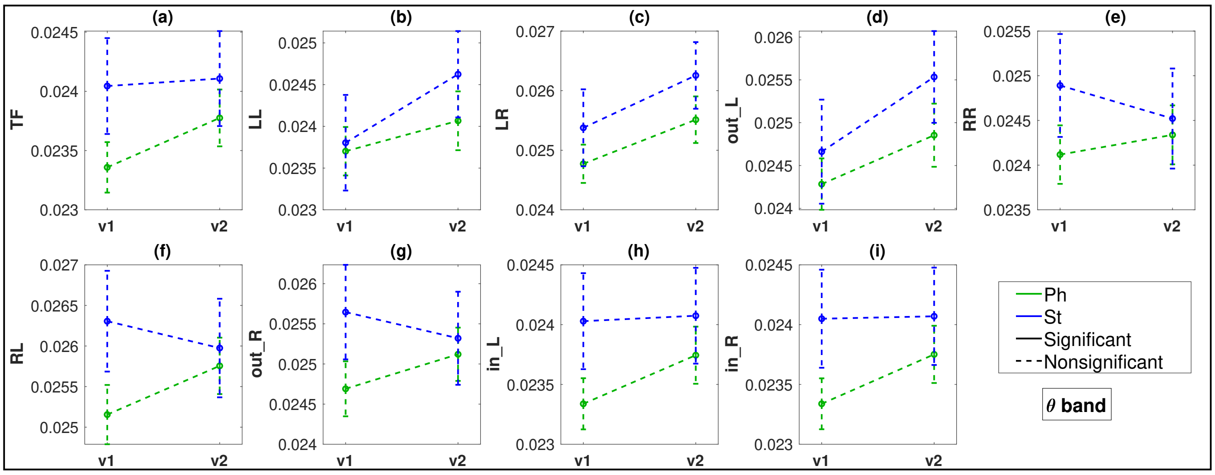


Figure 2: $\theta$ band: FLOW Metric comparisons between visits 1 (v1) and 2 (v2) for the pharmacological (Ph) and neurostimulation (St) groups. Metrics with $p < 0.05$ plotted in bold lines and $p \geq 0.05$ are in dotted lines.

Table 3: Repeated measure ANOVA OUTFLOW per channel metric results. Significant p values less than 0.05 are in bold font.

| | Fp1 | F3 | C3 | P3 | O1 | F7 | T3 | T5 | Fp2 | F4 | C4 | P4 | O2 | F8 | T4 | T6 | Fz | Cz | Pz |
|---|---|---|---|---|---|---|---|---|---|---|---|---|---|---|---|---|---|---|---|
| **RESPONSE** | | | | | | | | | | | | | | | | | | | |
| $\delta$ | 0.943 | 0.386 | 0.152 | 0.615 | 0.560 | 0.752 | 0.419 | 0.600 | 0.634 | 0.745 | 0.598 | 0.982 | 0.905 | 0.974 | 0.864 | **0.017** | 0.766 | 0.417 | 0.547 |
| $\theta$ | 0.775 | 0.861 | **0.045** | 0.242 | 0.598 | 0.451 | 0.066 | 0.427 | 0.846 | 0.869 | 0.651 | 0.576 | 0.781 | 0.712 | 0.977 | 0.402 | 0.085 | 0.529 | 0.942 |
| $\alpha$ | 0.748 | 0.668 | 0.970 | 0.176 | 0.192 | 0.547 | 0.597 | 0.520 | 0.898 | 0.691 | 0.103 | 0.718 | 0.328 | 0.478 | **0.037** | 0.055 | 0.166 | 0.251 | 0.515 |
| $\beta_1$ | 0.700 | 0.977 | 0.975 | 0.664 | 0.614 | 0.950 | 0.562 | **0.047** | 0.342 | 0.894 | 0.264 | 0.331 | 0.086 | 0.717 | 0.320 | 0.459 | 0.929 | 0.787 | 0.967 |
| $\beta_2$ | 0.213 | 0.696 | 0.938 | 0.661 | 0.556 | 0.829 | 0.736 | 0.106 | 0.188 | 0.397 | 0.914 | 0.779 | 0.311 | 0.360 | 0.951 | 0.890 | 0.854 | 0.518 | 0.974 |
| **TREATMENT** | | | | | | | | | | | | | | | | | | | |
| $\delta$ | 0.293 | 0.587 | **1.48E-04** | 0.936 | 0.188 | 0.389 | 0.336 | **0.048** | 0.142 | 0.418 | 0.205 | 0.972 | 0.576 | 0.590 | **0.042** | **0.021** | 0.641 | 0.689 | 0.136 |
| $\theta$ | 0.436 | 0.598 | 0.067 | 0.681 | 0.648 | 0.540 | 0.952 | **0.023** | 0.578 | 0.745 | 0.949 | 0.561 | **0.025** | 0.336 | 0.776 | 0.052 | 0.079 | 0.337 | 0.404 |
| $\alpha$ | 0.219 | 0.485 | 0.472 | 0.922 | 0.560 | 0.617 | 0.502 | 0.336 | 0.131 | 0.628 | 0.449 | **0.004** | 0.678 | 0.251 | 0.437 | 0.981 | 0.861 | 0.953 | 0.094 |
| $\beta_1$ | 0.558 | 0.701 | 0.979 | 0.912 | 0.152 | 0.130 | 0.074 | 0.632 | 0.681 | 0.418 | 0.451 | **0.011** | **1.55E-04** | 0.097 | 0.258 | 0.563 | 0.123 | 0.874 | 0.897 |
| $\beta_2$ | 0.836 | 0.832 | 0.319 | 0.830 | 0.874 | **0.021** | 0.258 | 0.119 | 0.330 | 0.332 | 0.628 | 0.064 | 0.100 | 0.070 | 0.191 | 0.901 | 0.786 | 0.379 | 0.638 |
| **VISIT** | | | | | | | | | | | | | | | | | | | |
| $\delta$ | 0.307 | 0.581 | 0.899 | 0.202 | 0.498 | **0.045** | 0.854 | 0.075 | 0.390 | 0.356 | 0.776 | 0.987 | 0.278 | 0.243 | 0.256 | 0.771 | 0.382 | 0.073 | 0.479 |
| $\theta$ | 0.127 | 0.599 | 0.577 | 0.051 | 0.060 | **0.047** | 0.577 | **0.002** | 0.479 | 0.383 | 0.803 | 0.900 | 0.430 | 0.635 | 0.739 | 0.442 | 0.356 | 0.240 | 0.889 |
| $\alpha$ | 0.343 | 0.606 | 0.885 | 0.905 | 0.060 | 0.543 | 0.646 | 0.511 | 0.555 | 0.574 | 0.679 | 0.225 | **0.036** | 0.968 | **0.040** | 0.318 | 0.485 | 0.693 | 0.217 |
| $\beta_1$ | 0.524 | 0.325 | **0.039** | 0.640 | 0.760 | 0.087 | 0.149 | 0.223 | 0.642 | 0.849 | 0.523 | 0.930 | 0.631 | 0.763 | 0.868 | 0.468 | 0.575 | 0.932 | 0.116 |
| $\beta_2$ | 0.635 | 0.708 | 0.245 | 0.810 | 0.877 | 0.243 | 0.344 | 0.405 | 0.595 | 0.917 | 0.205 | 0.577 | 0.514 | 0.397 | 0.075 | 0.590 | 0.163 | 0.368 | 0.121 |
| **RESPONSE:TREATMENT** | | | | | | | | | | | | | | | | | | | |
| $\delta$ | 0.341 | 0.654 | **0.008** | 0.249 | **0.003** | 0.982 | 0.438 | 0.246 | 0.322 | 0.109 | 0.690 | 0.194 | 0.546 | 0.386 | 0.296 | 0.282 | 0.225 | 0.265 | 0.785 |
| $\theta$ | 0.200 | 0.345 | **0.043** | 0.052 | 0.999 | 0.254 | 0.184 | 0.878 | 0.342 | 0.298 | 0.925 | 0.318 | 0.902 | 0.653 | 0.415 | 0.693 | 0.065 | 0.182 | 0.800 |
| $\alpha$ | 0.526 | 0.806 | 0.388 | 0.208 | 0.496 | 0.820 | 0.244 | 0.951 | 0.633 | 0.585 | 0.947 | 0.414 | 0.630 | 0.804 | 0.749 | 0.707 | 0.291 | 0.663 | 0.290 |
| $\beta_1$ | 0.126 | 0.351 | 0.988 | 0.365 | 0.187 | 0.663 | 0.617 | 0.197 | 0.817 | 0.651 | 0.871 | 0.737 | 0.718 | 0.068 | 0.442 | 0.799 | 0.856 | 0.182 | 0.950 |
| $\beta_2$ | 0.055 | 0.186 | 0.674 | 0.725 | 0.811 | 0.996 | 0.540 | 0.788 | 0.719 | 0.306 | 0.375 | 0.124 | 0.912 | 0.068 | 0.063 | 0.316 | 0.534 | 0.182 | 0.827 |
| **RESPONSE:VISIT** | | | | | | | | | | | | | | | | | | | |
| $\delta$ | 0.925 | 0.209 | 0.748 | 0.221 | 0.630 | 0.850 | 0.539 | 0.977 | 0.782 | 0.630 | 0.234 | 0.697 | 0.883 | 0.655 | 0.420 | 0.565 | 0.095 | 0.374 | **0.031** |
| $\theta$ | 0.479 | 0.409 | 0.977 | 0.433 | 0.443 | 0.378 | 0.444 | 0.871 | 0.681 | 0.227 | 0.493 | 0.739 | 0.797 | 0.063 | 0.464 | 0.467 | 0.243 | 0.341 | 0.813 |
| $\alpha$ | 0.704 | 0.389 | **0.035** | 0.905 | 0.426 | 0.774 | 0.072 | 0.695 | 0.280 | 0.438 | 0.951 | 0.591 | 0.245 | 0.894 | 0.201 | 0.750 | 0.141 | 0.733 | 0.551 |
| $\beta_1$ | 0.656 | 0.326 | 0.202 | 0.569 | 0.756 | 0.669 | 0.802 | 0.764 | 0.710 | 0.440 | 0.851 | 0.875 | 0.181 | **0.012** | 0.215 | 0.517 | 0.306 | 0.444 | 0.107 |
| $\beta_2$ | 0.888 | 0.412 | 0.659 | 0.972 | 0.984 | 0.772 | 0.274 | 0.439 | 0.964 | 0.518 | 0.512 | 0.953 | 0.144 | 0.064 | 0.739 | 0.352 | 0.671 | 0.891 | 0.537 |
| **TREATMENT:VISIT** | | | | | | | | | | | | | | | | | | | |
| $\delta$ | 0.368 | 0.086 | 0.102 | 0.404 | 0.730 | 0.964 | 0.353 | 0.244 | 0.338 | 0.477 | 0.202 | 0.689 | 0.928 | 0.745 | 0.828 | 0.926 | 0.426 | 0.866 | 0.158 |
| $\theta$ | 0.568 | 0.287 | 0.899 | 0.176 | 0.498 | 0.812 | 0.475 | 0.226 | 0.526 | 0.607 | 0.647 | 0.680 | 0.755 | 0.854 | 0.443 | 0.792 | 0.630 | 0.798 | 0.176 |
| $\alpha$ | 0.536 | 0.093 | 0.581 | 0.320 | 0.133 | 0.276 | 0.060 | 0.262 | 0.636 | 0.776 | 0.512 | 0.470 | 0.362 | 0.281 | 0.312 | 0.501 | **0.013** | 0.839 | 0.333 |
| $\beta_1$ | 0.827 | 0.395 | 0.950 | 0.228 | 0.786 | 0.517 | 0.980 | 0.648 | 0.480 | 0.112 | 0.362 | 0.422 | 0.726 | 0.111 | 0.216 | 0.504 | 0.060 | 0.057 | 0.805 |
| $\beta_2$ | 0.135 | **0.031** | 0.334 | 0.060 | 0.617 | 0.789 | 0.211 | 0.279 | 0.315 | 0.305 | 0.644 | 0.473 | 0.697 | 0.652 | 0.823 | 0.215 | 0.742 | 0.451 | 0.674 |
| **RESPONSE:TREATMENT:VISIT** | | | | | | | | | | | | | | | | | | | |
| $\delta$ | 0.978 | 0.981 | 0.575 | 0.460 | 0.974 | 0.213 | 0.748 | 0.595 | 0.564 | 0.709 | 0.725 | 0.737 | 0.713 | 0.191 | 0.865 | 0.532 | 0.687 | 0.299 | 0.261 |
| $\theta$ | 0.563 | 0.132 | 0.882 | 0.704 | 0.275 | 0.531 | 0.667 | 0.966 | 0.943 | 0.062 | 0.393 | 0.706 | 0.764 | 0.557 | 0.318 | 0.758 | 0.525 | 0.619 | 0.736 |
| $\alpha$ | 0.340 | 0.978 | 0.792 | 0.727 | 0.861 | 0.673 | 0.627 | 0.325 | 0.481 | 0.127 | 0.540 | 0.369 | 0.568 | 0.947 | 0.598 | 0.799 | 0.449 | 0.986 | 0.781 |
| $\beta_1$ | 0.309 | 0.129 | 0.191 | 0.549 | 0.958 | 0.120 | 0.656 | 0.685 | 0.654 | 0.092 | 0.962 | 0.540 | 0.965 | 0.513 | 0.333 | 0.462 | 0.532 | 0.289 | 0.780 |
| $\beta_2$ | 0.961 | 0.290 | 0.248 | 0.510 | 0.409 | 0.258 | 0.830 | 0.737 | 0.641 | 0.220 | 0.667 | 0.287 | 0.229 | 0.323 | 0.427 | 0.180 | 0.531 | 0.508 | 0.635 |

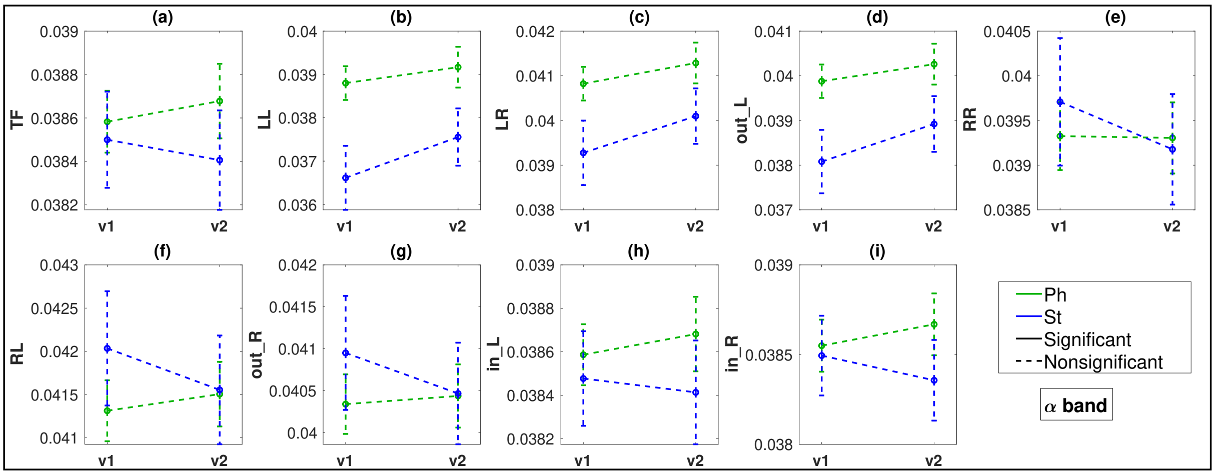


Figure 3: $\alpha$ band: FLOW Metric comparisons between visits 1 (v1) and 2 (v2) for the pharmacological (Ph) and neurostimulation (St) groups. Metrics with $p < 0.05$ plotted in bold lines and $p \geq 0.05$ are in dotted lines.

Table 4: **t-test, FDR adjusted t-test, Signrank** p-values for the comparison of **FLOW metrics** in **visit 1** vs **visit 2** across different EEG bands for pharmacological and neurostimulation separately. Significant p values less than 0.05 are in bold font.

| | **TF** | **LL** | **LR** | **out_L** | **RR** | **RL** | **out_R** | **in_L** | **in_R** |
|---|---|---|---|---|---|---|---|---|---|
| | | | | | **t-test** | | | | |
| | | | | | **Pharmacological: visit 1 vs visit 2** | | | | |
| $\delta$ | 0.809 | 0.931 | 0.610 | 0.747 | 0.918 | 0.444 | 0.712 | 0.732 | 0.912 |
| $\theta$ | **0.020** | 0.277 | **0.036** | 0.091 | 0.500 | 0.085 | 0.196 | **0.021** | **0.021** |
| $\alpha$ | 0.478 | 0.448 | 0.314 | 0.411 | 0.960 | 0.600 | 0.796 | 0.474 | 0.386 |
| $\beta_1$ | 0.744 | 0.446 | 0.504 | 0.510 | 0.101 | 0.081 | 0.082 | 0.598 | 0.967 |
| $\beta_2$ | **<0.001** | **0.043** | **0.016** | **0.024** | **0.010** | **0.049** | **0.021** | **<0.001** | **<0.001** |
| | | | | | **Neurostimulation: visit 1 vs visit 2** | | | | |
| $\delta$ | **0.004** | **0.001** | **0.002** | **<0.001** | 0.231 | 0.551 | 0.453 | **0.005** | **0.007** |
| $\theta$ | 0.748 | 0.073 | 0.056 | 0.053 | 0.457 | 0.512 | 0.516 | 0.815 | 0.920 |
| $\alpha$ | 0.536 | 0.128 | 0.118 | 0.129 | 0.370 | 0.390 | 0.394 | 0.691 | 0.394 |
| $\beta_1$ | 0.766 | 0.508 | 0.376 | 0.384 | 0.241 | 0.425 | 0.317 | 0.700 | 0.994 |
| $\beta_2$ | **0.040** | 0.106 | 0.205 | 0.127 | 0.277 | 0.287 | 0.291 | 0.100 | 0.056 |
| | | | | | **FDR adjusted t-test** | | | | |
| | | | | | **Pharmacological: visit 1 vs visit 2** | | | | |
| $\delta$ | 0.931 | 0.931 | 0.931 | 0.931 | 0.931 | 0.931 | 0.931 | 0.931 | 0.931 |
| $\theta$ | 0.064 | 0.312 | 0.082 | 0.136 | 0.500 | 0.136 | 0.251 | 0.064 | 0.064 |
| $\alpha$ | 0.718 | 0.718 | 0.718 | 0.718 | 0.960 | 0.772 | 0.895 | 0.718 | 0.718 |
| $\beta_1$ | 0.837 | 0.765 | 0.765 | 0.765 | 0.302 | 0.302 | 0.302 | 0.769 | 0.967 |
| $\beta_2$ | **<0.001** | **0.049** | **0.029** | **0.031** | **0.023** | **0.0497** | **0.031** | **<0.001** | **<0.001** |
| | | | | | **Neurostimulation: visit 1 vs visit 2** | | | | |
| $\delta$ | **0.009** | **0.002** | **0.006** | **0.002** | 0.297 | 0.551 | 0.510 | **0.010** | **0.010** |
| $\theta$ | 0.916 | 0.218 | 0.218 | 0.218 | 0.773 | 0.773 | 0.773 | 0.916 | 0.920 |
| $\alpha$ | 0.603 | 0.386 | 0.386 | 0.386 | 0.507 | 0.507 | 0.507 | 0.691 | 0.507 |
| $\beta_1$ | 0.862 | 0.761 | 0.761 | 0.761 | 0.761 | 0.761 | 0.761 | 0.862 | 0.994 |
| $\beta_2$ | 0.229 | 0.229 | 0.291 | 0.229 | 0.291 | 0.291 | 0.291 | 0.229 | 0.229 |
| | | | | | **Signrank** | | | | |
| | | | | | **Pharmacological: visit 1 vs visit 2** | | | | |
| $\delta$ | 0.528 | 0.842 | 0.885 | 0.734 | 0.822 | 0.588 | 0.911 | 0.772 | 0.498 |
| $\theta$ | **0.016** | 0.428 | 0.080 | 0.170 | 0.565 | 0.152 | 0.291 | **0.029** | **0.014** |
| $\alpha$ | 0.820 | 0.590 | 0.480 | 0.580 | 0.539 | 0.919 | 0.820 | 0.930 | 0.999 |
| $\beta_1$ | 0.529 | 0.372 | 0.540 | 0.467 | 0.139 | 0.131 | 0.118 | 0.428 | 0.741 |
| $\beta_2$ | **0.000** | 0.067 | **0.018** | **0.033** | **0.005** | **0.022** | **0.009** | **0.0005** | **0.000** |
| | | | | | **Neurostimulation: visit 1 vs visit 2** | | | | |
| $\delta$ | **0.007** | **0.002** | **0.005** | **0.001** | 0.330 | 0.582 | 0.478 | **0.007** | **0.017** |
| $\theta$ | 0.711 | 0.063 | 0.063 | 0.045 | 0.325 | 0.386 | 0.374 | 0.832 | 0.975 |
| $\alpha$ | 0.427 | 0.061 | 0.052 | 0.067 | 0.336 | 0.228 | 0.295 | 0.519 | 0.380 |
| $\beta_1$ | 0.816 | 0.539 | 0.446 | 0.485 | 0.397 | 0.634 | 0.519 | 0.619 | 0.767 |
| $\beta_2$ | 0.085 | 0.258 | 0.512 | 0.336 | 0.176 | 0.162 | 0.197 | 0.220 | 0.101 |

Table 5: **t-test, FDR adjusted t-test, Signrank** p-values for the comparison of **INFLOW per channel** in **visit 1** vs **visit 2** across different EEG bands for pharmacological and neurostimulation separately. Significant p values less than 0.05 are in bold font.

| | Fp1 | F3 | C3 | P3 | O1 | F7 | T3 | T5 | Fp2 | F4 | C4 | P4 | O2 | F8 | T4 | T6 | Fz | Cz | Pz |
|---|---|---|---|---|---|---|---|---|---|---|---|---|---|---|---|---|---|---|---|
| **t-test** | | | | | | | | | | | | | | | | | | | |
| **Pharmacological: visit 1 vs visit 2** | | | | | | | | | | | | | | | | | | | |
| $\delta$ | 0.563 | 0.505 | 0.697 | 0.513 | 0.936 | 0.317 | 0.678 | 0.775 | 0.316 | 0.787 | 0.644 | 0.927 | 0.963 | 0.843 | 0.320 | 0.682 | 0.664 | 0.729 | 0.998 |
| $\theta$ | 0.067 | **0.047** | **0.028** | **0.015** | **0.020** | **0.033** | 0.061 | **0.006** | 0.052 | **0.014** | **0.033** | **0.050** | **0.018** | **0.031** | **0.037** | **0.014** | **0.048** | **0.013** | **0.015** |
| $\alpha$ | 0.479 | 0.419 | 0.642 | 0.411 | 0.425 | 0.602 | 0.516 | 0.993 | 0.289 | 0.324 | 0.585 | 0.411 | 0.457 | 0.310 | 0.538 | 0.899 | 0.763 | 0.827 | 0.957 |
| $\beta_1$ | 0.516 | 0.865 | 0.599 | 0.264 | 0.970 | 1.000 | 0.774 | 0.454 | 0.927 | 0.265 | 0.466 | 0.602 | 0.771 | 0.141 | 0.759 | 0.428 | 0.533 | 0.545 | 0.550 |
| $\beta_2$ | 0.200 | **<0.001** | **0.009** | **<0.001** | **<0.001** | **0.041** | 0.375 | **0.005** | **<0.001** | **0.002** | **<0.001** | **<0.001** | **<0.001** | **0.005** | 0.494 | **<0.001** | **<0.001** | **<0.001** | **<0.001** |
| **Neurostimulation: visit 1 vs visit 2** | | | | | | | | | | | | | | | | | | | |
| $\delta$ | **0.010** | **0.013** | **0.040** | **0.017** | 0.100 | 0.114 | **0.025** | **0.002** | 0.121 | **0.023** | 0.104 | **0.009** | **0.010** | 0.124 | 0.054 | **0.002** | **0.016** | **0.002** | **0.005** |
| $\theta$ | 0.980 | 0.322 | 0.677 | 0.844 | 0.717 | 0.838 | 0.192 | 0.971 | 0.569 | 0.606 | 0.903 | 0.323 | 0.469 | 0.981 | 0.596 | 0.870 | 0.121 | 0.543 | 0.387 |
| $\alpha$ | 0.196 | 0.861 | 0.745 | 0.735 | 0.797 | **0.048** | 0.694 | 0.473 | 0.303 | 0.709 | 0.582 | 0.277 | 0.946 | 0.271 | 0.670 | 0.188 | 0.749 | 0.437 | 0.209 |
| $\beta_1$ | 0.359 | 0.716 | 0.071 | 0.844 | 0.718 | 0.185 | 0.564 | 0.856 | 0.985 | 0.324 | 0.516 | 0.713 | 0.581 | 0.704 | 0.870 | 0.755 | 0.945 | 0.194 | 0.565 |
| $\beta_2$ | 0.687 | 0.709 | 0.827 | 0.050 | **0.008** | 0.646 | 0.369 | 0.210 | 0.161 | 0.206 | 0.114 | 0.915 | **0.048** | 0.217 | 0.200 | 0.729 | **0.012** | 0.082 | 0.177 |
| **FDR adjusted t-test** | | | | | | | | | | | | | | | | | | | |
| **Pharmacological: visit 1 vs visit 2** | | | | | | | | | | | | | | | | | | | |
| $\delta$ | 0.998 | 0.998 | 0.998 | 0.998 | 0.998 | 0.998 | 0.998 | 0.998 | 0.998 | 0.998 | 0.998 | 0.998 | 0.998 | 0.998 | 0.998 | 0.998 | 0.998 | 0.998 | 0.998 |
| $\theta$ | 0.067 | 0.058 | 0.052 | **0.048** | **0.048** | 0.052 | 0.065 | **0.048** | 0.058 | **0.048** | 0.052 | 0.058 | **0.048** | 0.052 | 0.054 | **0.048** | 0.058 | **0.048** | **0.048** |
| $\alpha$ | 0.871 | 0.871 | 0.871 | 0.871 | 0.871 | 0.871 | 0.871 | 0.993 | 0.871 | 0.871 | 0.871 | 0.871 | 0.871 | 0.871 | 0.871 | 0.993 | 0.967 | 0.982 | 0.993 |
| $\beta_1$ | 0.953 | 1.000 | 0.953 | 0.953 | 1.000 | 1.000 | 0.980 | 0.953 | 1.000 | 0.953 | 0.953 | 0.953 | 0.980 | 0.953 | 0.980 | 0.953 | 0.953 | 0.953 | 0.953 |
| $\beta_2$ | 0.224 | **0.001** | **0.012** | **<0.001** | **<0.001** | **0.049** | 0.396 | **0.007** | **<0.001** | **0.002** | **0.001** | **<0.001** | **<0.001** | **0.007** | 0.494 | **<0.001** | **<0.001** | **0.002** | **0.001** |
| **Neurostimulation: visit 1 vs visit 2** | | | | | | | | | | | | | | | | | | | |
| $\delta$ | **0.028** | **0.032** | 0.058 | **0.032** | 0.124 | 0.124 | **0.040** | **0.013** | 0.124 | **0.040** | 0.124 | **0.028** | **0.028** | 0.124 | 0.073 | **0.013** | **0.032** | **0.013** | **0.023** |
| $\theta$ | 0.981 | 0.981 | 0.981 | 0.981 | 0.981 | 0.981 | 0.981 | 0.981 | 0.981 | 0.981 | 0.981 | 0.981 | 0.981 | 0.981 | 0.981 | 0.981 | 0.981 | 0.981 | 0.981 |
| $\alpha$ | 0.823 | 0.909 | 0.890 | 0.890 | 0.890 | 0.823 | 0.890 | 0.890 | 0.823 | 0.890 | 0.890 | 0.823 | 0.946 | 0.823 | 0.890 | 0.823 | 0.890 | 0.890 | 0.823 |
| $\beta_1$ | 0.973 | 0.973 | 0.973 | 0.973 | 0.973 | 0.973 | 0.973 | 0.973 | 0.985 | 0.973 | 0.973 | 0.973 | 0.973 | 0.973 | 0.973 | 0.973 | 0.985 | 0.973 | 0.973 |
| $\beta_2$ | 0.815 | 0.815 | 0.873 | 0.240 | 0.114 | 0.815 | 0.539 | 0.344 | 0.344 | 0.344 | 0.344 | 0.915 | 0.240 | 0.344 | 0.344 | 0.815 | 0.114 | 0.312 | 0.344 |
| **Signrank** | | | | | | | | | | | | | | | | | | | |
| **Pharmacological: visit 1 vs visit 2** | | | | | | | | | | | | | | | | | | | |
| $\delta$ | 0.852 | 0.634 | 0.824 | 0.898 | 0.661 | 0.218 | 0.911 | 0.954 | **0.068** | 0.978 | 0.616 | 0.661 | 0.788 | 0.386 | 0.646 | 0.932 | 0.915 | 0.588 | 0.436 |
| $\theta$ | **0.076** | **0.043** | **0.044** | **0.038** | **0.031** | **0.039** | 0.167 | **0.005** | 0.051 | **0.016** | **0.029** | 0.089 | **0.016** | **0.017** | 0.054 | **0.008** | 0.052 | **0.010** | **0.011** |
| $\alpha$ | 0.626 | 0.950 | 0.883 | 0.757 | 0.542 | 0.881 | 0.603 | 0.784 | 0.581 | 0.764 | 0.795 | 0.906 | 0.761 | 0.907 | 0.689 | 0.779 | 0.948 | 0.606 | 0.520 |
| $\beta_1$ | 0.502 | 0.699 | 0.337 | 0.120 | 0.902 | 0.975 | 0.832 | 0.477 | 0.892 | 0.193 | 0.318 | 0.499 | 0.547 | 0.206 | 0.738 | 0.207 | 0.460 | 0.317 | 0.644 |
| $\beta_2$ | 0.158 | **0.000** | **0.005** | **0.000** | **0.000** | **0.013** | 0.565 | **0.004** | **0.000** | **0.000** | **0.000** | **0.000** | **0.000** | **0.004** | 0.799 | **0.000** | **0.000** | **0.000** | **0.000** |
| **Neurostimulation: visit 1 vs visit 2** | | | | | | | | | | | | | | | | | | | |
| $\delta$ | **0.013** | **0.020** | **0.018** | 0.054 | 0.101 | 0.186 | 0.053 | **0.004** | 0.112 | **0.041** | 0.186 | **0.025** | **0.026** | 0.092 | 0.075 | **0.007** | **0.015** | **0.007** | **0.011** |
| $\theta$ | 0.899 | 0.427 | 0.759 | 0.882 | 0.403 | 0.975 | 0.179 | 0.975 | 0.695 | 0.924 | 0.907 | 0.519 | 0.409 | 0.966 | 0.512 | 0.866 | 0.179 | 0.589 | 0.346 |
| $\alpha$ | 0.156 | 0.933 | 0.664 | 0.582 | 0.899 | 0.081 | 0.539 | 0.440 | 0.285 | 0.519 | 0.743 | 0.110 | 0.975 | 0.216 | 0.857 | 0.204 | 0.561 | 0.649 | 0.346 |
| $\beta_1$ | 0.325 | 0.634 | **0.048** | 0.767 | 0.525 | 0.427 | 0.374 | 0.949 | 0.568 | 0.459 | 0.446 | 0.492 | 0.519 | 0.397 | 0.512 | 0.589 | 0.849 | 0.212 | 0.711 |
| $\beta_2$ | 0.657 | 0.310 | 0.759 | 0.156 | **0.009** | 0.363 | 0.276 | 0.267 | 0.117 | 0.179 | 0.120 | 0.767 | 0.112 | 0.159 | 0.153 | 0.891 | **0.038** | **0.083** | 0.244 |

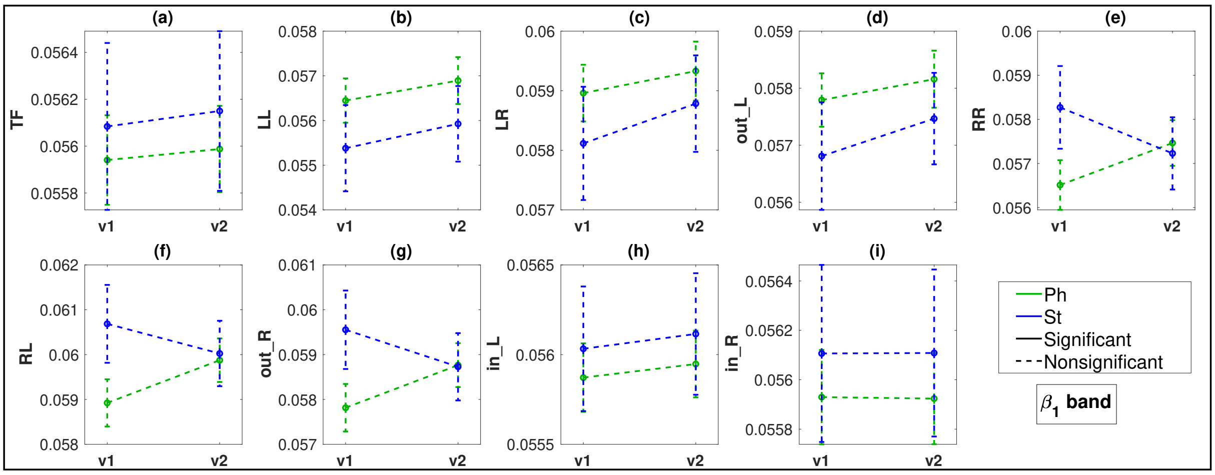


Figure 4: $\beta_1$ band: FLOW Metric comparisons between visits 1 (v1) and 2 (v2) for the pharmacological (Ph) and neurostimulation (St) groups. Metrics with $p < 0.05$ plotted in bold lines and $p \geq 0.05$ are in dotted lines.

Table 6: **t-test, FDR adjusted t-test, ranksum** p-values for the comparison of the **FLOW metrics** at **visit 1** between **respondents (R)** and **nonrespondents (NR)** for the two treatment groups. Significant p-values less than 0.05 are in bold font.

| | **TF** | **LL** | **LR** | **out_L** | **RR** | **RL** | **out_R** | **in_L** | **in_R** |
|---|---|---|---|---|---|---|---|---|---|
| **t-test** | | | | | | | | | |
| **Pharmacological: R visit 1 vs NR visit 1** | | | | | | | | | |
| $\boldsymbol{\delta}$ | 0.642 | 0.703 | 0.353 | 0.637 | 0.478 | 0.346 | 0.953 | 0.580 | 0.697 |
| $\boldsymbol{\theta}$ | 0.288 | **0.026** | **0.044** | **0.038** | 0.876 | 0.799 | 0.767 | 0.344 | 0.279 |
| $\boldsymbol{\alpha}$ | **0.020** | 0.818 | 0.535 | 0.675 | **0.022** | **0.026** | **0.021** | **0.027** | **0.018** |
| $\boldsymbol{\beta_1}$ | 0.119 | 0.267 | 0.292 | 0.271 | 0.214 | 0.184 | 0.169 | 0.107 | 0.127 |
| $\boldsymbol{\beta_2}$ | 0.387 | 0.974 | 0.749 | 0.860 | 0.609 | 0.542 | 0.591 | 0.291 | 0.472 |
| **Neurostimulation: R visit 1 vs NR visit 1** | | | | | | | | | |
| $\boldsymbol{\delta}$ | **0.037** | 0.531 | 0.343 | 0.465 | 0.364 | 0.288 | 0.197 | **0.026** | **0.041** |
| $\boldsymbol{\theta}$ | 0.236 | 0.256 | 0.200 | 0.205 | 0.944 | 0.911 | 0.910 | 0.261 | 0.212 |
| $\boldsymbol{\alpha}$ | 0.680 | 0.368 | 0.317 | 0.362 | 0.568 | 0.640 | 0.652 | 0.777 | 0.574 |
| $\boldsymbol{\beta_1}$ | 0.749 | 0.071 | **0.041** | 0.053 | 0.140 | 0.265 | 0.204 | 0.883 | 0.756 |
| $\boldsymbol{\beta_2}$ | 0.130 | 0.120 | 0.052 | 0.095 | **0.032** | 0.066 | 0.052 | 0.062 | 0.244 |
| **FDR adjusted t-test** | | | | | | | | | |
| **Pharmacological: R visit 1 vs NR visit 1** | | | | | | | | | |
| $\boldsymbol{\delta}$ | 0.790 | 0.790 | 0.790 | 0.790 | 0.790 | 0.790 | 0.953 | 0.790 | 0.790 |
| $\boldsymbol{\theta}$ | 0.516 | 0.131 | 0.131 | 0.131 | 0.876 | 0.876 | 0.876 | 0.516 | 0.516 |
| $\boldsymbol{\alpha}$ | **0.040** | 0.818 | 0.688 | 0.760 | **0.040** | **0.040** | **0.040** | **0.040** | **0.040** |
| $\boldsymbol{\beta_1}$ | 0.292 | 0.292 | 0.292 | 0.292 | 0.292 | 0.292 | 0.292 | 0.292 | 0.292 |
| $\boldsymbol{\beta_2}$ | 0.914 | 0.974 | 0.963 | 0.968 | 0.914 | 0.914 | 0.914 | 0.914 | 0.914 |
| **Neurostimulation: R visit 1 vs NR visit 1** | | | | | | | | | |
| $\boldsymbol{\delta}$ | 0.123 | 0.531 | 0.468 | 0.523 | 0.468 | 0.468 | 0.443 | 0.123 | 0.123 |
| $\boldsymbol{\theta}$ | 0.391 | 0.391 | 0.391 | 0.391 | 0.944 | 0.944 | 0.944 | 0.391 | 0.391 |
| $\boldsymbol{\alpha}$ | 0.765 | 0.765 | 0.765 | 0.765 | 0.765 | 0.765 | 0.765 | 0.777 | 0.765 |
| $\boldsymbol{\beta_1}$ | 0.851 | 0.212 | 0.212 | 0.212 | 0.316 | 0.398 | 0.367 | 0.883 | 0.851 |
| $\boldsymbol{\beta_2}$ | 0.146 | 0.146 | 0.119 | 0.142 | 0.119 | 0.119 | 0.119 | 0.119 | 0.244 |
| **ranksum** | | | | | | | | | |
| **Pharmacological: R visit 1 vs NR visit 1** | | | | | | | | | |
| $\boldsymbol{\delta}$ | 0.563 | 0.747 | 0.452 | 0.656 | 0.570 | 0.312 | 0.934 | 0.452 | 0.586 |
| $\boldsymbol{\theta}$ | 0.257 | **0.024** | **0.036** | **0.034** | 0.676 | 0.697 | 0.596 | 0.309 | 0.237 |
| $\boldsymbol{\alpha}$ | 0.057 | 0.938 | 0.757 | 0.901 | **0.017** | **0.040** | **0.020** | 0.058 | 0.050 |
| $\boldsymbol{\beta_1}$ | 0.205 | 0.234 | 0.251 | 0.237 | 0.307 | 0.277 | 0.267 | 0.189 | 0.207 |
| $\boldsymbol{\beta_2}$ | 0.823 | 0.622 | 0.374 | 0.504 | 0.529 | 0.609 | 0.599 | 0.583 | 0.845 |
| **Neurostimulation: R visit 1 vs NR visit 1** | | | | | | | | | |
| $\boldsymbol{\delta}$ | 0.074 | 0.508 | 0.307 | 0.479 | 0.552 | 0.174 | 0.119 | **0.049** | 0.082 |
| $\boldsymbol{\theta}$ | 0.387 | 0.694 | 0.425 | 0.425 | 0.849 | 0.866 | 0.849 | 0.375 | 0.375 |
| $\boldsymbol{\alpha}$ | 0.466 | 0.329 | 0.412 | 0.351 | 0.955 | 0.831 | 0.884 | 0.508 | 0.552 |
| $\boldsymbol{\beta_1}$ | 0.814 | 0.055 | **0.047** | **0.049** | 0.182 | 0.307 | 0.266 | 0.849 | 0.814 |
| $\boldsymbol{\beta_2}$ | 0.307 | 0.113 | 0.082 | 0.099 | **0.047** | 0.082 | 0.064 | 0.113 | 0.400 |

Table 7: **t-test, FDR adjusted t-test, ranksum** p-values for the comparison of the **INFLOW per channel metrics** at **visit 1** between **respondents (R)** and **nonrespondents (NR)** for the two treatment groups. Significant p-values less than 0.05 are in bold font.

| | **Fp1** | **F3** | **C3** | **P3** | **O1** | **F7** | **T3** | **T5** | **Fp2** | **F4** | **C4** | **P4** | **O2** | **F8** | **T4** | **T6** | **Fz** | **Cz** | **Pz** |
|---|---|---|---|---|---|---|---|---|---|---|---|---|---|---|---|---|---|---|---|
| t-test | | | | | | | | | | | | | | | | | | | |
| **Pharmacological: R visit 1 vs NR visit 1** | | | | | | | | | | | | | | | | | | | |
| $\delta$ | 0.560 | 0.683 | 0.298 | 0.408 | 0.604 | 0.621 | 0.648 | 0.614 | 0.574 | 0.687 | 0.386 | 0.542 | 0.663 | 0.630 | 0.988 | 0.276 | 0.932 | 0.685 | 0.456 |
| $\theta$ | 0.423 | 0.336 | 0.287 | 0.452 | 0.252 | 0.479 | 0.381 | 0.275 | 0.314 | 0.304 | 0.219 | 0.315 | 0.326 | 0.353 | 0.279 | 0.228 | 0.244 | 0.251 | 0.130 |
| $\alpha$ | **0.015** | 0.062 | **0.033** | **0.035** | 0.113 | **0.003** | 0.311 | 0.061 | **0.028** | **0.013** | **0.034** | **0.007** | **0.008** | **0.030** | 0.325 | 0.083 | **0.034** | 0.068 | **0.010** |
| $\beta_1$ | **0.035** | 0.223 | 0.366 | 0.080 | 0.312 | 0.232 | 0.064 | **0.034** | 0.071 | 0.431 | 0.142 | 0.179 | 0.172 | 0.220 | 0.152 | 0.051 | 0.324 | 0.215 | 0.074 |
| $\beta_2$ | 0.199 | 0.169 | 0.351 | 0.930 | 0.650 | 0.176 | 0.479 | 0.695 | 0.160 | 0.351 | 0.931 | 0.590 | 0.990 | 0.261 | 0.809 | 0.810 | 0.410 | 0.790 | 0.500 |
| **Neurostimulation: R visit 1 vs NR visit 1** | | | | | | | | | | | | | | | | | | | |
| $\delta$ | **0.012** | 0.099 | 0.081 | 0.121 | 0.122 | 0.061 | **0.022** | **0.019** | 0.068 | 0.371 | **0.004** | 0.059 | **0.039** | 0.113 | 0.095 | 0.100 | 0.134 | 0.118 | 0.133 |
| $\theta$ | 0.258 | 0.164 | 0.223 | 0.400 | 0.224 | 0.304 | 0.220 | 0.412 | 0.182 | 0.180 | 0.177 | 0.172 | 0.281 | 0.253 | 0.273 | 0.254 | 0.410 | 0.305 | 0.114 |
| $\alpha$ | 0.808 | 0.645 | 0.925 | 0.362 | 0.770 | 0.794 | 0.445 | 0.905 | 0.434 | 0.564 | 0.896 | 0.620 | 0.583 | 0.234 | 0.820 | 0.939 | 0.501 | 0.665 | 0.526 |
| $\beta_1$ | 0.893 | 0.854 | 0.699 | 0.967 | 0.835 | 0.906 | 0.625 | 0.863 | 0.807 | 0.961 | 0.915 | 0.782 | 0.515 | 0.461 | 0.944 | 0.610 | 0.618 | 0.412 | 0.404 |
| $\beta_2$ | 0.363 | 0.134 | 0.405 | 0.183 | 0.338 | **0.041** | **0.018** | 0.447 | 0.289 | **0.020** | 0.098 | 0.546 | 0.291 | 0.452 | 0.741 | 0.296 | 0.297 | 0.249 | 0.254 |
| FDR adjusted t-test | | | | | | | | | | | | | | | | | | | |
| **Pharmacological: R visit 1 vs NR visit 1** | | | | | | | | | | | | | | | | | | | |
| $\delta$ | 0.767 | 0.767 | 0.767 | 0.767 | 0.767 | 0.767 | 0.767 | 0.767 | 0.767 | 0.767 | 0.767 | 0.767 | 0.767 | 0.767 | 0.988 | 0.767 | 0.984 | 0.767 | 0.767 |
| $\theta$ | 0.473 | 0.447 | 0.447 | 0.478 | 0.447 | 0.479 | 0.453 | 0.447 | 0.447 | 0.447 | 0.447 | 0.447 | 0.447 | 0.447 | 0.447 | 0.447 | 0.447 | 0.447 | 0.447 |
| $\alpha$ | **0.048** | 0.084 | 0.055 | 0.055 | 0.126 | **0.048** | 0.325 | 0.084 | 0.055 | **0.048** | 0.055 | **0.048** | **0.048** | 0.055 | 0.325 | 0.099 | 0.055 | 0.086 | **0.048** |
| $\beta_1$ | 0.218 | 0.294 | 0.387 | 0.218 | 0.362 | 0.294 | 0.218 | 0.218 | 0.218 | 0.431 | 0.294 | 0.294 | 0.294 | 0.294 | 0.294 | 0.218 | 0.362 | 0.294 | 0.218 |
| $\beta_2$ | 0.943 | 0.943 | 0.943 | 0.983 | 0.961 | 0.943 | 0.943 | 0.961 | 0.943 | 0.943 | 0.983 | 0.961 | 0.994 | 0.943 | 0.961 | 0.961 | 0.943 | 0.961 | 0.943 |
| **Neurostimulation: R visit 1 vs NR visit 1** | | | | | | | | | | | | | | | | | | | |
| $\delta$ | 0.106 | 0.142 | 0.142 | 0.142 | 0.142 | 0.142 | 0.106 | 0.106 | 0.142 | 0.371 | 0.082 | 0.142 | 0.142 | 0.142 | 0.142 | 0.142 | 0.142 | 0.142 | 0.142 |
| $\theta$ | 0.363 | 0.363 | 0.363 | 0.412 | 0.363 | 0.363 | 0.363 | 0.412 | 0.363 | 0.363 | 0.363 | 0.363 | 0.363 | 0.363 | 0.363 | 0.363 | 0.412 | 0.363 | 0.363 |
| $\alpha$ | 0.939 | 0.939 | 0.939 | 0.939 | 0.939 | 0.939 | 0.939 | 0.939 | 0.939 | 0.939 | 0.939 | 0.939 | 0.939 | 0.939 | 0.939 | 0.939 | 0.939 | 0.939 | 0.939 |
| $\beta_1$ | 0.967 | 0.967 | 0.967 | 0.967 | 0.967 | 0.967 | 0.967 | 0.967 | 0.967 | 0.967 | 0.967 | 0.967 | 0.967 | 0.967 | 0.967 | 0.967 | 0.967 | 0.967 | 0.967 |
| $\beta_2$ | 0.492 | 0.471 | 0.505 | 0.471 | 0.492 | 0.259 | 0.192 | 0.505 | 0.471 | 0.192 | 0.465 | 0.576 | 0.471 | 0.505 | 0.741 | 0.471 | 0.471 | 0.471 | 0.471 |
| ranksum | | | | | | | | | | | | | | | | | | | |
| **Pharmacological: R visit 1 vs NR visit 1** | | | | | | | | | | | | | | | | | | | |
| $\delta$ | 0.510 | 0.964 | 0.232 | 0.323 | 0.690 | 0.583 | 0.768 | 0.718 | 0.548 | 0.976 | 0.405 | 0.478 | 0.535 | 0.612 | 0.957 | 0.259 | 0.972 | 0.639 | 0.277 |
| $\theta$ | 0.305 | 0.269 | 0.298 | 0.321 | 0.316 | 0.335 | 0.267 | 0.277 | 0.335 | 0.257 | 0.178 | 0.267 | 0.219 | 0.305 | 0.228 | 0.204 | 0.228 | 0.184 | 0.110 |
| $\alpha$ | **0.033** | 0.129 | 0.065 | 0.073 | 0.307 | **0.012** | 0.364 | 0.117 | 0.056 | **0.028** | 0.094 | **0.012** | **0.027** | 0.063 | 0.427 | 0.171 | **0.049** | 0.133 | **0.014** |
| $\beta_1$ | **0.048** | 0.307 | 0.478 | 0.192 | 0.548 | 0.366 | 0.073 | **0.041** | 0.118 | 0.570 | 0.155 | 0.253 | 0.408 | 0.344 | 0.143 | 0.087 | 0.410 | 0.523 | 0.122 |
| $\beta_2$ | 0.239 | 0.366 | 0.199 | 0.540 | 0.940 | 0.314 | 0.649 | 0.972 | 0.287 | 0.455 | 0.711 | 0.740 | 0.660 | 0.279 | 0.871 | 0.890 | 0.870 | 0.878 | 0.870 |
| **Neurostimulation: R visit 1 vs NR visit 1** | | | | | | | | | | | | | | | | | | | |
| $\delta$ | **0.017** | 0.135 | 0.174 | 0.297 | 0.104 | 0.148 | 0.052 | 0.052 | 0.119 | 0.413 | **0.005** | 0.099 | 0.055 | 0.167 | 0.329 | 0.124 | 0.154 | 0.130 | 0.197 |
| $\theta$ | 0.425 | 0.297 | 0.318 | 0.479 | 0.375 | 0.567 | 0.425 | 0.645 | 0.297 | 0.425 | 0.257 | 0.307 | 0.522 | 0.363 | 0.413 | 0.508 | 0.745 | 0.582 | 0.197 |
| $\alpha$ | 0.920 | 0.537 | 0.955 | 0.266 | 0.425 | 0.728 | 0.257 | 0.552 | 0.452 | 0.678 | 0.902 | 0.902 | 0.425 | 0.307 | 0.920 | 0.745 | 0.375 | 0.678 | 0.363 |
| $\beta_1$ | 0.973 | 0.955 | 0.866 | 0.920 | 0.991 | 0.866 | 0.613 | 0.866 | 0.866 | 0.884 | 0.884 | 0.866 | 0.598 | 0.522 | 0.711 | 0.494 | 0.582 | 0.413 | 0.508 |
| $\beta_2$ | 0.413 | 0.351 | 0.425 | 0.375 | 0.413 | 0.119 | **0.006** | 0.522 | 0.351 | **0.030** | 0.104 | 0.694 | 0.318 | 0.387 | 0.779 | 0.182 | 0.466 | 0.479 | 0.329 |

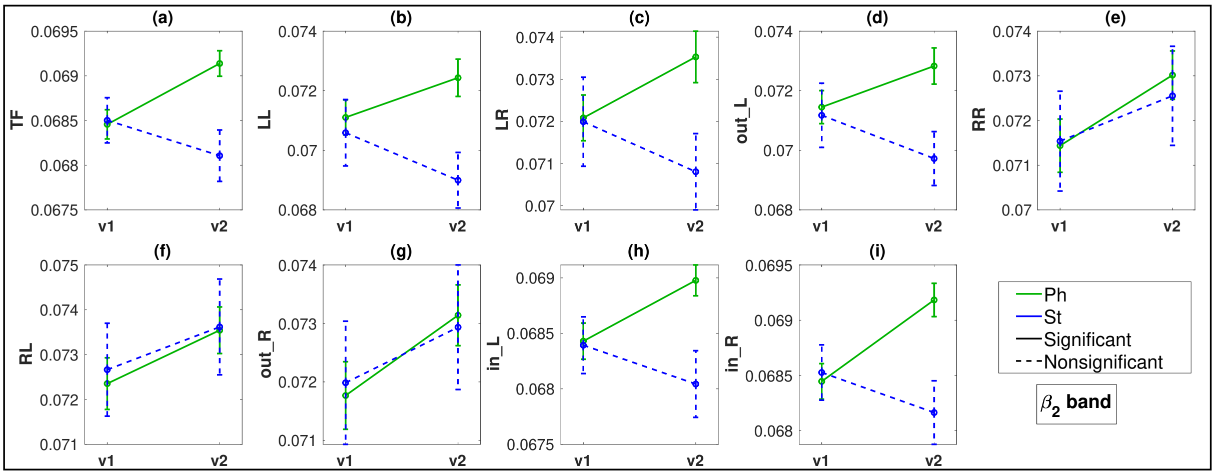


Figure 5: $\beta_2$ band: FLOW Metric comparisons between visits 1 (v1) and 2 (v2) for the pharmacological (Ph) and neurostimulation (St) groups. Metrics with $p < 0.05$ plotted in bold lines and $p \geq 0.05$ are in dotted lines.

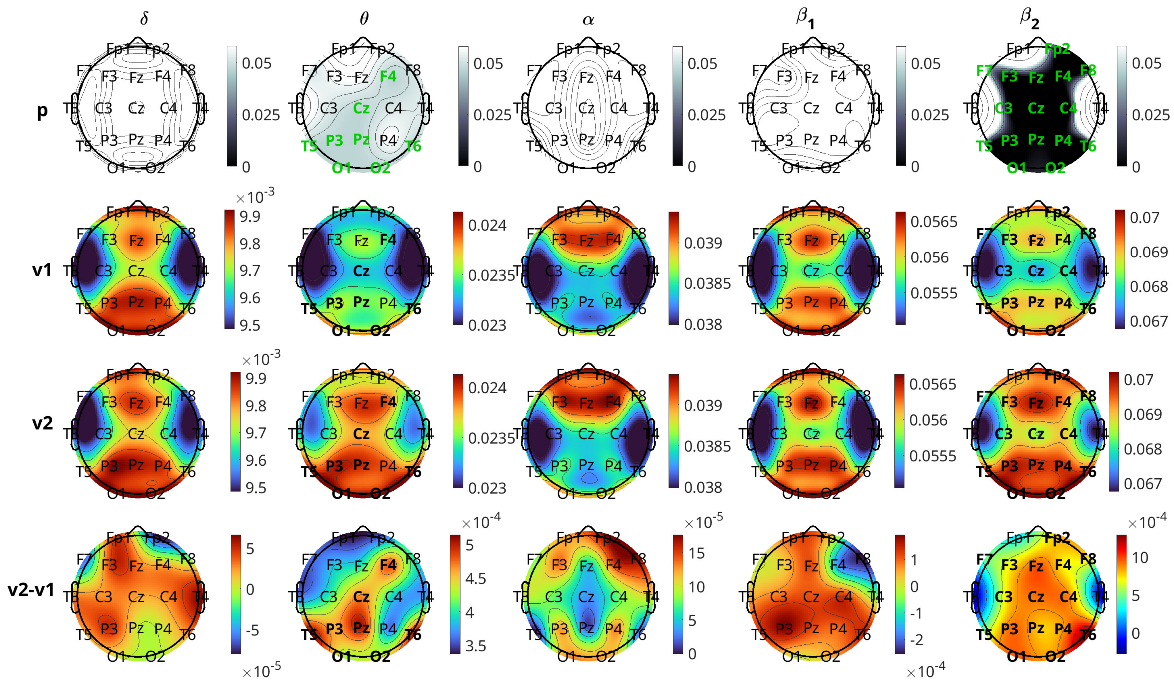

Figure 6: Brain topographical plots for the comparison of inflow per channel across the two visits 1 (v1) and 2 (v2), for the pharmacological group. The first row of panels is the p-values for the comparison. The 2nd and 3rd rows present the average values (across subjects) of the inflow for each visit, while the 4th row presents the difference. Columns correspond to frequency bands, as indicated by the text above the top panels. Channels with p-values $< 0.05$ are indicated in green-coloured text in the first row.

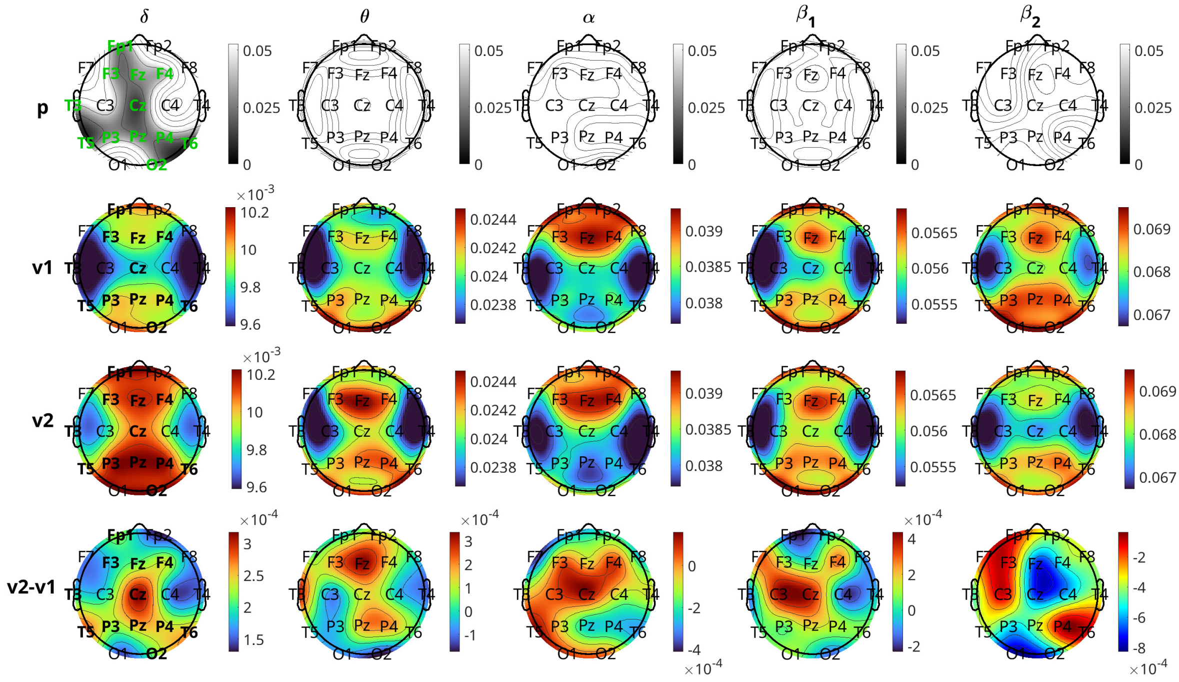

Figure 7: Brain topographical plots for the comparison of inflow per channel across the two visits 1 (v1) and 2 (v2), for the neurostimulation group. The first row of panels is the p-values for the comparison. The 2nd and 3rd rows present the average values (across subjects) of the inflow for each visit, while the 4th row presents the difference. Columns correspond to frequency bands, as indicated by the text above the top panels. Channels with p-values $< 0.05$ are indicated in green-coloured text in the first row.

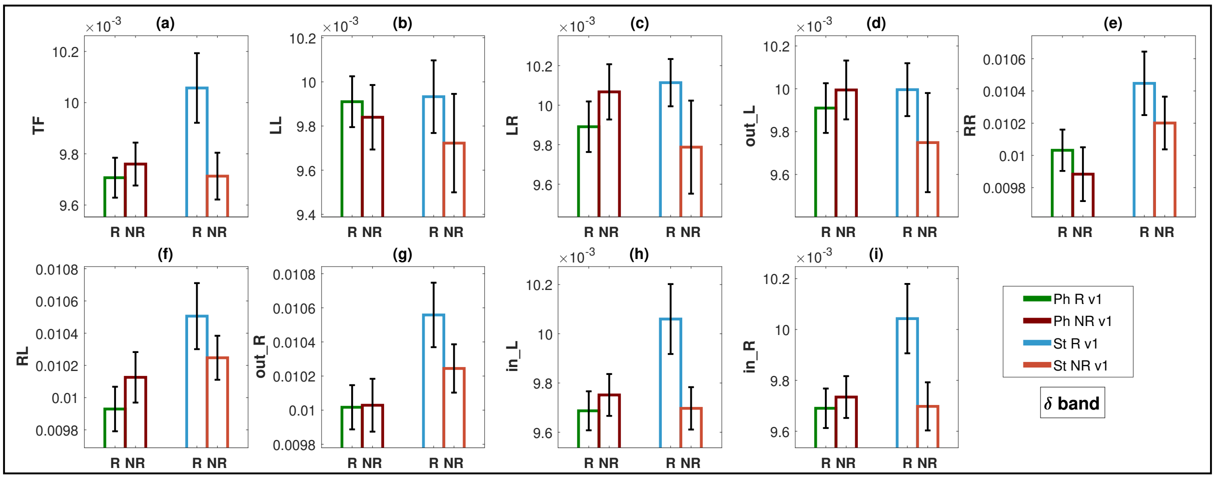


Figure 8: $\delta$ band: Bar plots representing the FLOW metrics at the time of visit 1 for respondents (R) and nonrespondents (NR) to pharmacological (Ph) and neurostimulation (St) treatment. Metrics with $p < 0.05$ have face colours and for $p \geq 0.05$ have no face colours in the bar plots. The first two bar plots in each panel are for Ph and the next two are for St.

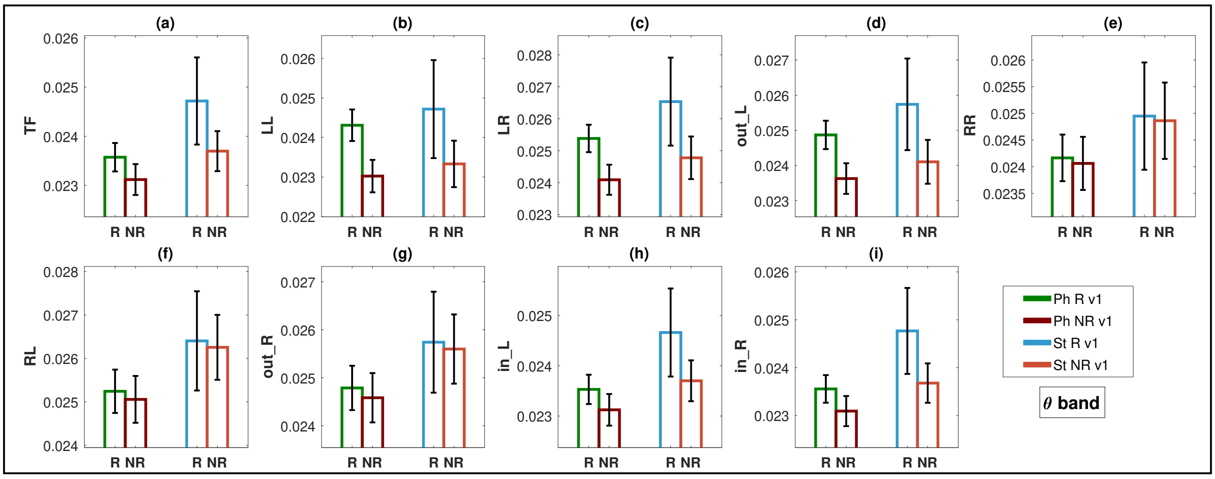


Figure 9: $\theta$ band: Bar plots representing the FLOW metrics at the time of visit 1 for respondents (R) and nonrespondents (NR) to pharmacological (Ph) and neurostimulation (St) treatment. Metrics with $p < 0.05$ have face colours and for $p \geq 0.05$ have no face colours in the bar plots. The first two bar plots in each panel are for Ph and the next two are for St.

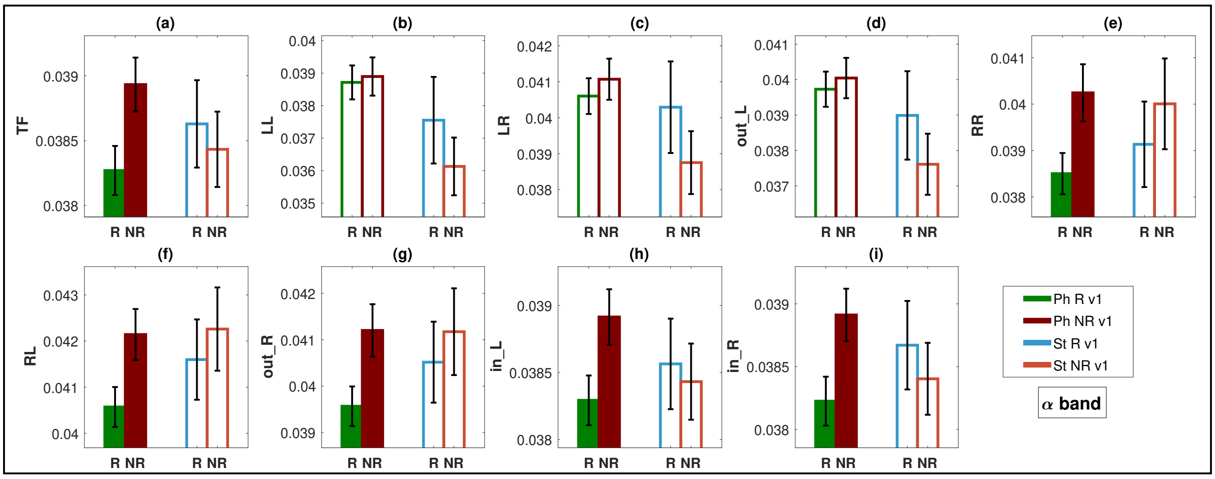


Figure 10: $\alpha$ band: Bar plots representing the FLOW metrics at the time of visit 1 for respondents (R) and nonrespondents (NR) to pharmacological (Ph) and neurostimulation (St) treatment. Metrics with $p < 0.05$ have face colours and for $p \geq 0.05$ have no face colours in the bar plots. The first two bar plots in each panel are for Ph and the next two are for St.

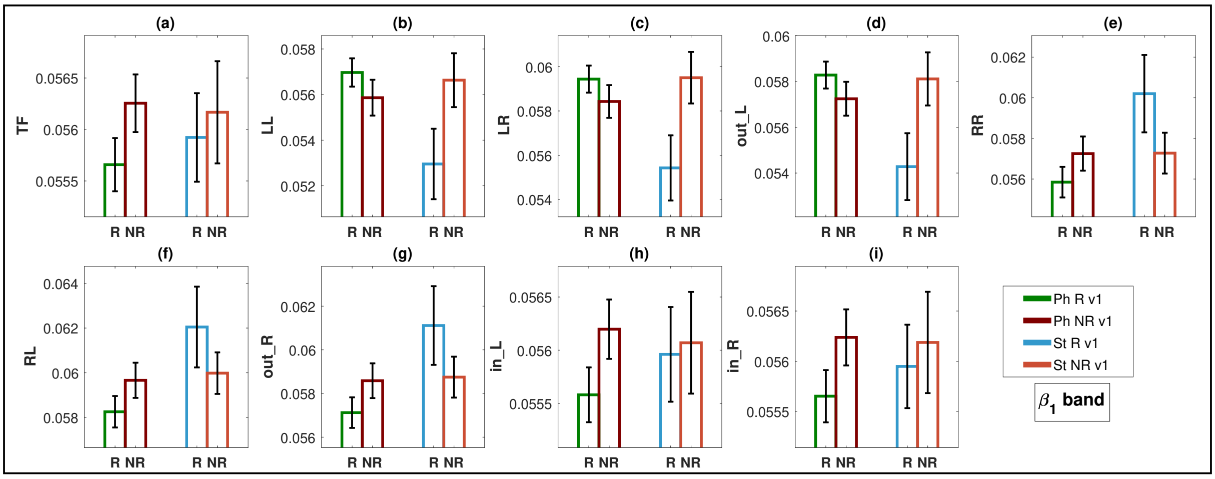


Figure 11: $\beta_1$ band: Bar plots representing the FLOW metrics at the time of visit 1 for respondents (R) and nonrespondents (NR) to pharmacological (Ph) and neurostimulation (St) treatment. Metrics with $p < 0.05$ have face colours and for $p \geq 0.05$ have no face colours in the bar plots. The first two bar plots in each panel are for Ph and the next two are for St.

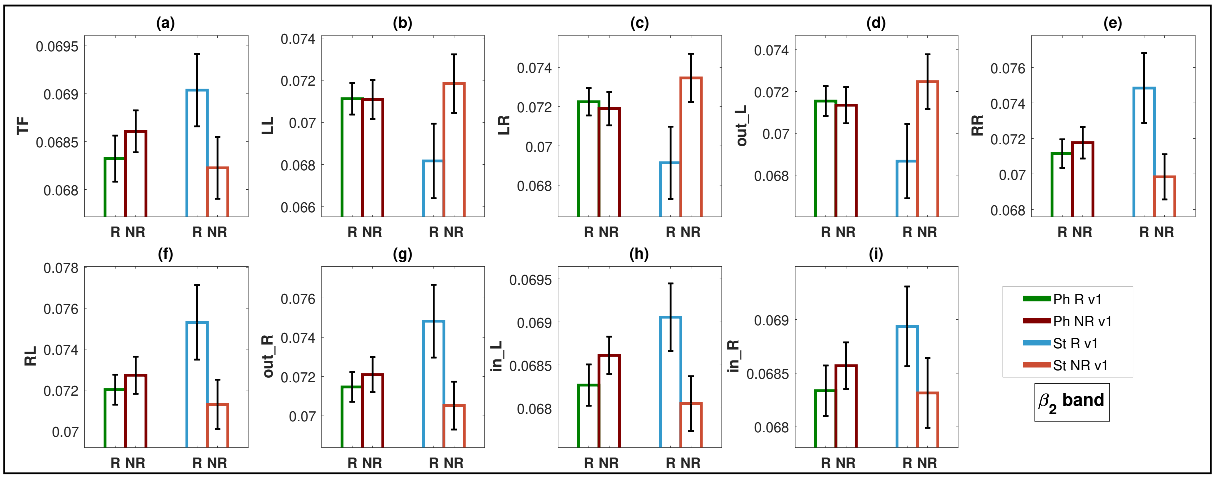


Figure 12: $\beta_2$ band: Bar plots representing the FLOW metrics at the time of visit 1 for respondents (R) and nonrespondents (NR) to pharmacological (Ph) and neurostimulation (St) treatment. Metrics with $p < 0.05$ have face colours and for $p \geq 0.05$ have no face colours in the bar plots. The first two bar plots in each panel are for Ph and the next two are for St.

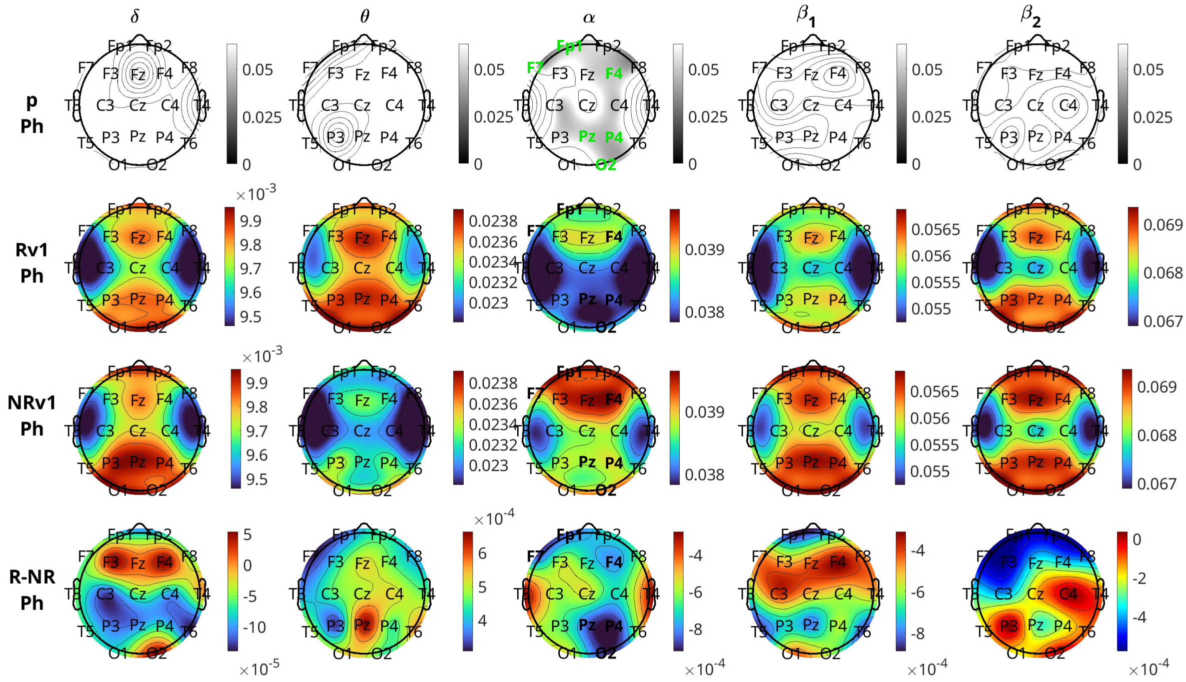


Figure 13: Brain topographical plots for the comparison of inflow per channel between respondents (R) and nonrespondents (NR) for the pharmacological (Ph) group at visit 1 (v1). The first row of panels is the p-values. The second and the third rows are the average inflow per channel (across all subjects) at visit 1 for respondents (R) and nonrespondents (NR). The fourth row is the difference in inflow between R and NR averaged over all subjects. Columns correspond to frequency bands, as indicated by the text above the top panels. Channels with p-values $< 0.05$ are indicated in green-coloured text in the 1st and 3rd rows.

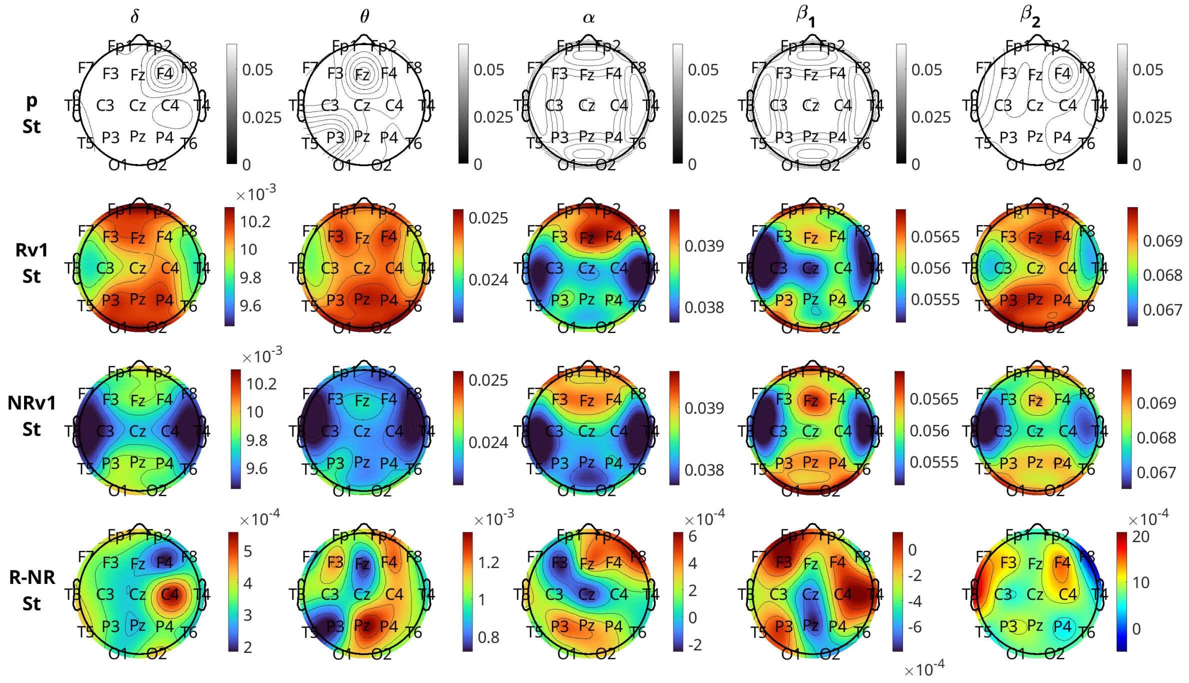

Figure 14: Brain topographical plots for the comparison of inflow per channel between respondents (R) and nonrespondents (NR) for the neurostimulation (St) group at visit 1 (v1). The first row of panels is the p-values. The second and the third rows are the average inflow per channel (across all subjects) at visit 1 for respondents (R) and nonrespondents (NR). The fourth row is the difference in inflow between R and NR averaged over all subjects. Columns correspond to frequency bands, as indicated by the text above the top panels. Channels with p-values $< 0.05$ are indicated in green-coloured text in the 1st and 3rd rows.
